# Mapping internet activity in Australian cities during COVID-19 lockdown: how occupational factors drive inequality

**DOI:** 10.1101/2021.02.04.21251171

**Authors:** Cameron Zachreson, Erika Martino, Martin Tomko, Freya M. Shearer, Rebecca Bentley, Nicholas Geard

## Abstract

During the COVID-19 pandemic, evidence has accumulated that movement restrictions enacted to combat virus spread produce disparate consequences along socioeconomic lines. We investigate the hypothesis that people engaged in financially secure employment are better able to adhere to mobility restrictions, due to occupational factors that link the capacity for flexible work arrangements to income security. We use high-resolution spatial data on household internet traffic as a surrogate for adaptation to home-based work, together with the geographical clustering of occupation types, to investigate the relationship between occupational factors and increased internet traffic during work hours under lockdown in two Australian cities. By testing our hypothesis based on the observed trends, and exploring demographic factors associated with divergences from our hypothesis, we are left with a picture of unequal impact dominated by two major influences: the types of occupations in which people are engaged, and the composition of households and families. During lockdown, increased internet traffic was correlated with income security and, when school activity was conducted remotely, to the proportion of families with children. Our findings suggest that response planning and provision of social and economic support for residents within lockdown areas should explicitly account for income security and household structure. Overall, the results we present contribute to the emerging picture of the impacts of COVID-19 on human behaviour, and will help policy makers to understand the balance between public health and social impact in making decisions about mitigation policies.

## I. INTRODUCTION

Globally, the COVID-19 pandemic has changed the ways in which people live and work. Governments have used various mitigation strategies to stem the spread of infection by restricting mobility and limiting the rate of physical interaction between people [1]. In Australia, these strategies have included limiting the size of gatherings, restricting travel across state borders, closing workplaces, and encouraging working and schooling from home. To help ensure compliance with these policies, and to dampen their short-term economic impact, the Australian government in early 2020 implemented social assistance programs, including the temporary COVID Supplementary payment for the unemployed (JobSeeker), the JobKeeper wage subsidy, early access to superannuation (retirement) accounts, and targeted support for severely affected sectors [2, 3]. However, these measures cannot be maintained indefinitely, and the protracted COVID-19 pandemic may outlast Australia’s ability to continue providing the high levels of economic support that helped ensure compliance with restrictions. The broad aim of the present study is to quantify and map socioeconomic factors that are likely to determine current and future adherence to mobility restrictions. Detailed knowledge of the geographic distribution of these factors will assist when predicting the social impact of targeted mitigation strategies during future outbreaks of COVID-19, or subsequent pandemics.

Broadly, we expect a reduced effectiveness and increased economic impact of such mobility restrictions where there is a concentration of people who cannot adapt to home-based work. We know, for example, that people in factory-based occupations, hospitality, and essential services are unable to work from home [4–7]. The confluence of occupational and financial constraints place many such workers at a greater risk of exposure to infectious disease, either through the occupational hazard of close social interactions, or because without adequate leave or income entitlements, they have a limited ability to remain at home when unwell [6, 8, 9]. This conflict between household economic needs and public health orders to stay at home is problematic for the success of such mitigation strategies.

COVID-19 has driven a general economic contraction brought about through concerted reductions in consumer, recreational, and occupational activity [10–12]. Impact disparities can be explained (at least partially) by examining the distribution of employment types within and between subpopulations. Those who can perform their work requirements at home from a computer with internet access have experienced less-severe economic impact [13–18]. By engaging in written communication, and replacing face-to-face interactions with online video conferencing, work-related tasks are likely to result in measurable increases to home internet traffic. Population-level data on home internet usage may therefore provide a useful complement to the widely available mobility data typically used to monitor and model the real-time effects of COVID-19 and the restrictions associated with mitigation measures [16, 17, 19– 27]. While mobility data can tell us who is staying home and where people are going when they leave the home, internet volume data provides a unique perspective on what is happening within households, particularly in relation to adapting work arrangements to COVID-19 lockdown requirements.

Here, we demonstrate how relationships between occupational factors and home internet traffic can provide insight into social and economic disparities that are amplified by the pandemic and associated mitigation strategies. Our results support the hypothesis that occupational factors link the ability to work from home with income security, and clearly show how this link produces strong positive correlations between income security and increased home internet activity during COVID-19 restrictions. Our results in the Australian context may help explain observations from other recent studies describing the connection between income, internet, and the ability to self-isolate during COVID-19 [17, 21]. Overall, the results we present contribute to the emerging picture of the impacts of COVID-19 on human behaviour, and will help policy makers to understand the balance between public health and social impact in making future decisions. Furthermore, results such as those presented in this work will contribute to the ability to produce precise, integrated models of epidemic dynamics connected to social and economic phenomena.

## II. METHODS

### A. Data sources

Our study examines the Greater Metropolitan Areas of Melbourne and Sydney, Australia. To subdivide these regions, the geographic analysis unit adopted was the ASGS Statistical Area Level 2 (SA2). SA2 regions are defined by the Australian Bureau of Statistics (ABS) and typically contain between 1,000 and 10,000 residents, representing neighbourhoods that socially and economically interact [28]. Aggregating to this geographic unit, we used the following data sets to create our measures of occupational factors, income security and internet usage. We leveraged detailed population surveys quantifying the distributions of occupation types within local regions, and the characteristics of different occupation classifications [29–32]. To quantify changes to household internet volume, we used a population-scale data set describing home internet use patterns aggregated on the scale of SA2. This data was obtained from nbn co ltd. (**nbn**^TM^), a Government Business Enterprise providing national wholesale broadband access in Australia. Finally, we used recent survey data collected in September, 2020 by the COVID-19 Attitudes Resilience and Epidemiology (CARE) study to substantiate our individual-level interpretation of observed population-scale trends [33].

### B. Computation of income security by occupation

To quantify the salient features of the complex distributions of employment characteristics in Sydney and Melbourne, we constructed an income security index using data on employment security and income characteristics linked to the Australian and New Zealand Standard Classification of Occupations (ANZSCO). For income, we used average weekly earnings by occupation from the ABS Census of 2016 [34]. To calculate the employment security associated with an occupation, we used the most recent iteration (2018) of the nationally-representative Household, Income and Labour Dynamics in Australia (HILDA) survey. For each occupation, we computed income security as the product of the proportion of securely employed HILDA respondents, and the average weekly wage reported by the ABS (with income rescaled to the sample maximum). This gives a value between 0 and 1, with zero corresponding to occupations with no securely employed individuals and values of 1 corresponding to occupations with maximal remuneration as well as 100% securely employed respondents. The distributions of these values and the component measures of income and proportion securely employed are shown in the Supporting Material Fig. S3.

Occupation security status was developed as an individual categorical variable with two levels: [0 (secure employment)] or [1 (insecure employment)]. An individual was classified as ‘secure’ if they had a fixed-term or permanent job. We computed the employment security score associated with each occupation as the proportion of respondents in each occupation who were securely employed. We then computed the index of income security by occupation as the product of the employment security score and average weekly earnings (rescaled to the sample maximum). Distributions of the resulting income security values and the component measures of income and proportion securely employed are shown in Fig. S3. Note that those on a fixed-term contract were classified as securely employed because the work conditions associated with fixed-term employment are more similar to the conditions of permanent employment than they are to casual work conditions. Those on fixed-term contracts have also been shown to be more sociodemographically similar to those employed permanently than to those employed on a casual basis [35, 36].

### C. Computation of ability to work from home by occupation

To compute the working from home indicator for each occupation classification, we adapted an analysis done by Dingel and Neiman [37] (available on GitHub, https://github.com/jdingel/DingelNeiman-workathome) to establish which occupations could potentially be performed from home. Dingel and Neiman used the ‘Work Context’ and ‘Generalized Work Activities’ occupational surveys from the O*NET® Database. Drawing on a series of questions, they classified occupations according to whether they were compatible with working from home. For example, occupations were considered to be unsuitable for working from home if they involved activities that required a workplace such as operation of machinery or handling of specialised items (for more information in methodology, please refer to Dingel [37]). To produce international estimates we linked the binary work-from-home classifications (N = 969) produced by Dingel and Nieman, to International Standard Classification of Occupations (ISCO-08) 4-digit codes. This data (N = 1024) was then linked to the Australian and New Zealand Standard Classification of Occupations (ANZSCO) 6-digit codes and subsequently aggregated to the level of ANZSCO 4-digit codes for compatibility with ABS occupation distributions by SA2. During this linking and aggregation process, some of the occupation codes did not link (N = 36). For these, binary values were manually determined by referring to similar occupation descriptions from the original O*Net and SOC descriptions.

### D. Income security and ability to work from home as SA2-level measures

To quantify the ability to work from home and income security (respectively) for each SA2, we computed the weighted average of the respective variables by occupation based on the occupation distribution in each region. Weights correspond to the fraction of employed individuals working in each occupation within each SA2 region as reported by the ABS Census of 2016. The distributions of employed persons by occupation (4-digit level), by SA2 region was generated using the Census TableBuilder application provided by the ABS [30]. The data set queried was the 2016 Census Count of Persons by Usual Residence, aggregated by OCCP 4-Digit Level, by SA2 (UR). Distributions of the resulting SA2-level values and the component measures are shown in Fig. S5.

### E. Analysis of internet usage data from nbn^TM^

**nbn**^TM^ provided access to aggregated Australian internet usage volume data from household customers. The data provided by **nbn**^TM^ consist of upload and download volume (in bytes) by individual households over 30 minute intervals, spatially aggregated into SA2 regions. Different types of internet usage (such as streaming movies, videoconferencing and online gaming) are associated with different patterns of upload and download volume. The high time resolution and structure of the data allowed us to approximately differentiate between background (latent) internet activity, and active internet use. The data were restricted to the total download and upload volume for 30min intervals, per SA2 region, and the corresponding number of active internet connections per time slot that generated these data. Only data generated by at least 50 domestic connections were provided, to avoid privacy concerns and to reduce the impact of aberrant individual household behaviours in regions with insufficient service coverage.

Outlier data points beyond three standard deviations from the corresponding time period mean were removed, to limit the impact of outlier usage data points, usually caused by network management or internal infrastructure configuration changes. We set the data collection interval of October 10th, 2019 until November 29th, 2019 as the baseline period, when life in Australia was not impacted by school holidays or major, longer public holidays, and preceding the major disturbance produced by the bushfire season of summer 2019-2020. The period representing behaviour during the 1st wave of COVID-19 restrictions was set to the interval of April 18th - April 24th, 2020, while the period representing the second wave of restrictions was set to the interval of August 8th - August 14th, 2020.

The following upload and download volume characteristics per SA2 region were computed for these three periods (baseline and the two COVID-19 intervals): (1) an overall daytime average volume; (2) the daily average minimum volume relating to the minimum internet usage, between 4:30am and 5:30am; (3) the average volume generated during the daytime period (from school start until noon, 9:00am - 12:00pm noon); and (4) the average daily maximum usage (8:30pm - 10:00pm). Figure 1 demonstrates the daily fluctuation of average internet use for the period of September 2019 - September 2020 in Australia.

**FIG. 1.**
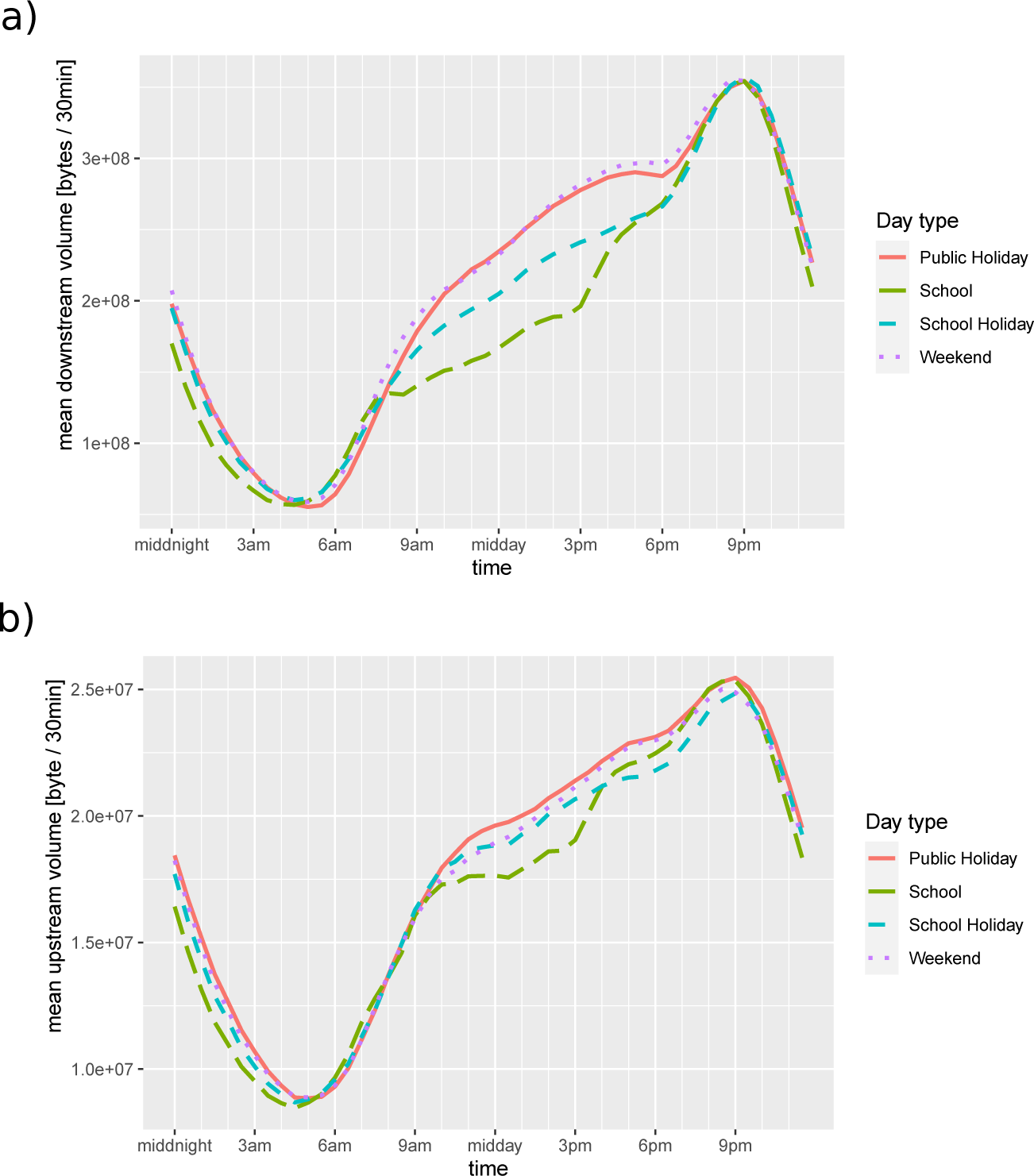
Long-term daily fluctuation of average internet activity per active service, per 30min period, for downloads (a), and uploads (b). The daily minimum and maximum, as well as the daytime plateau of activity are clearly visible. The changes between normal work days (school days), weekends and public holidays are salient. The daily trends in this figure were aggregated over the period from Sep. 1st 2019, to Sep. 1st 2020.

### F. Censoring of outlier data for correlation analysis

All correlation coefficients were computed after censoring data points for which either variable was greater than three standard deviations from the sample mean. The full data set made available with this article contains all values including outlier data. Outlier data is not included in the scatter plots shown in Figure 5 and Figure 6.

**FIG. 5.**
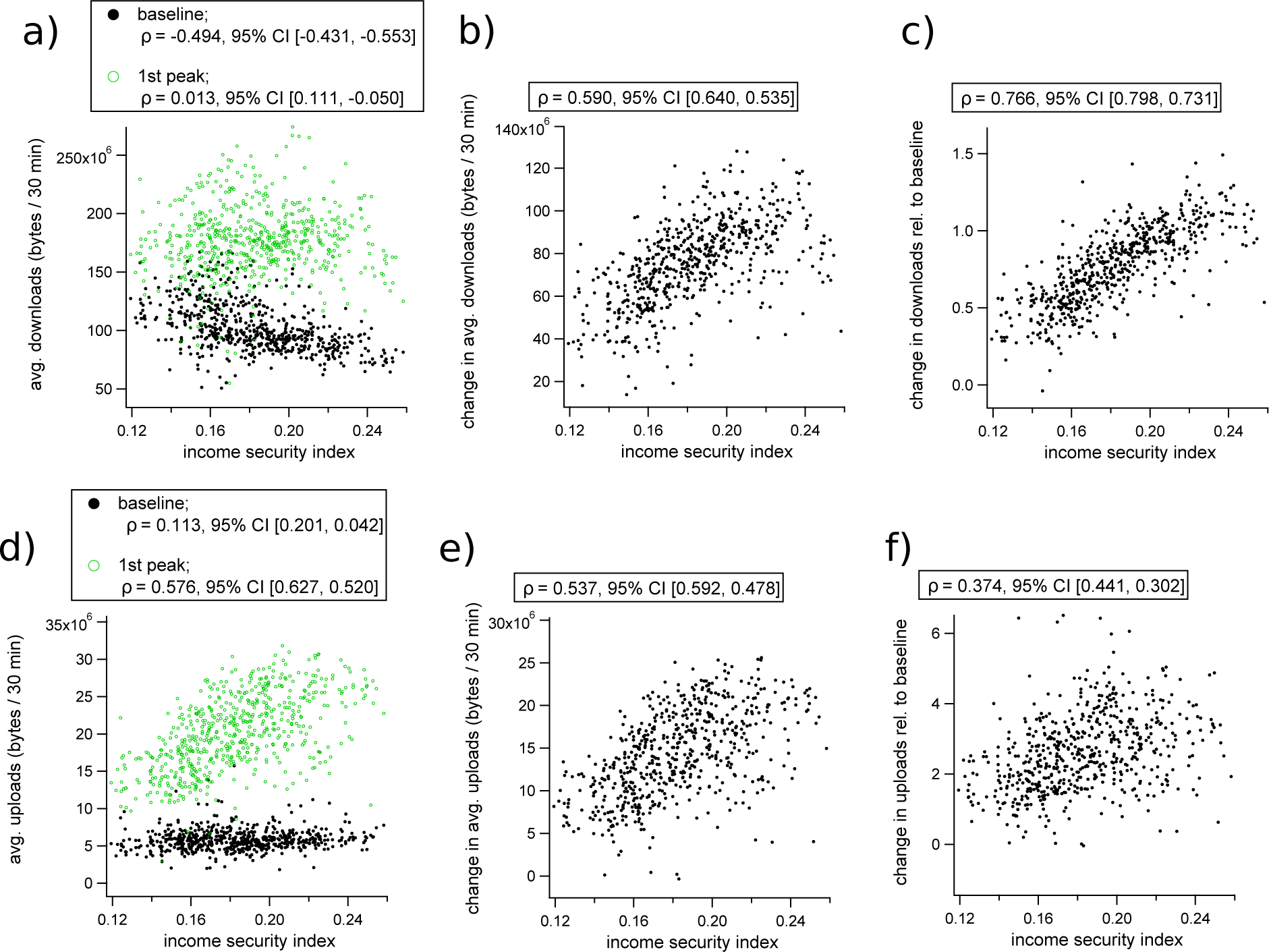
Changes to internet use during the 1st wave of COVID-19 restrictions in Australia, plotted against income security for each SA2 region. (a) Shows absolute average household download rates before (black dots) and during (green dots) the selected period (April 18th to April 24th, 2020). (b) Plots the absolute change in average per-household download rate during the first-wave period, and (c) plots the change in download rate relative to baseline. (d) Shows absolute upload rates before (black) and during (green) COVID-19, while (e) shows absolute changes to upload volume, and (f) shows changes to upload volume relative to baseline. In each subplot, the internet traffic quantifiers are plotted against the income security score for the corresponding SA2 region, and Pearson’s correlation coefficients with 95% CI intervals are shown in the legends.

**FIG. 6.**
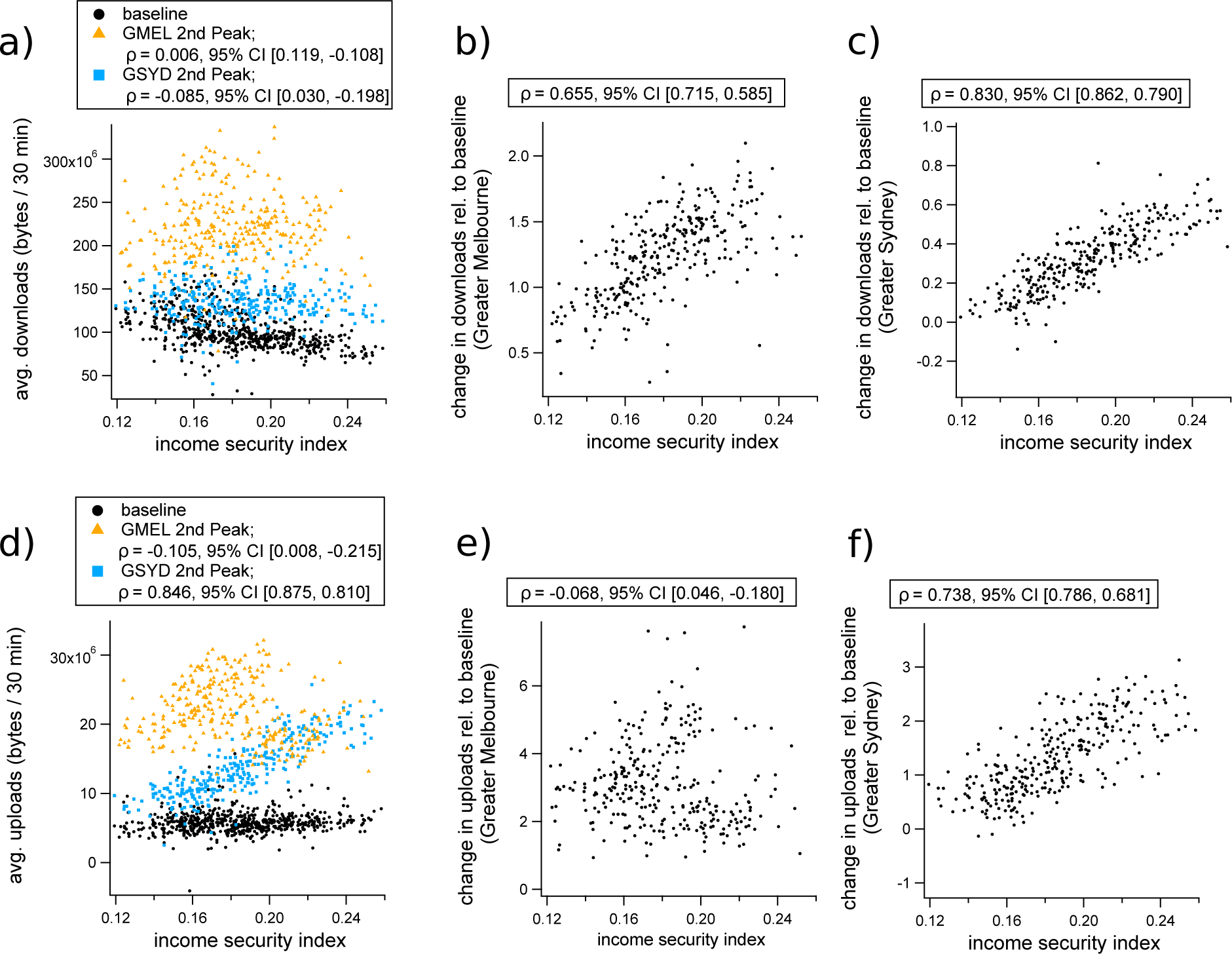
Changes to internet use during the second wave of COVID-19 restrictions in Australia, plotted against income security for each SA2 region. (a) Shows absolute download volumes averaged over the baseline period and second wave of COVID-19 restriction policies (August 8th to August 14th, 2020), which includes baseline (black dots), 2nd wave values for Greater Sydney (blue squares), and second wave values for Greater Melbourne (orange triangles). Plots (a) and (c) show the change in download volumes relative to pre-COVID baseline as a function of income security index for each SA2 in Sydney and Melbourne, respectively. Plots (d), (e) and (f) show the same analysis as (a), (b), and (c), respectively, for uploads rather than downloads. In each subplot, the internet traffic quantifiers are plotted against the income security score for the corresponding SA2 region, and Pearson’s correlation coefficients with 95% CI intervals are shown in the legends.

### G. Analysis of data from the CARE survey

Our analysis used data from the CARE study’s Victoria-wide survey which aimed to address the overall question: How were Victorians thinking, feeling and behaving in response to the ‘second wave’ of the COVID-19 epidemic and the associated public health measures. The survey was self-administered online in English to 1006 Victorian residents aged 18 years and over. The survey was based on research developed and conducted by Imperial College in the UK in mid-March 2020 [33, 38]. Some questions in the Australian survey were modified slightly to reflect local response measures and terminology. Additional questions were added to the Australian survey to measure social and emotional impacts. Data collection in both the UK and Australia was conducted by the online market research agency YouGov.

The CARE study used a structured questionnaire addressing the following three domains: perceptions of risk and consequences of COVID-19 infection; measures taken by individuals to protect themselves and others from COVID-19 infection; and social and emotional impact. The questionnaire was administered online to members of the YouGov Australia panel of individuals who have agreed to take part in surveys of public opinion (over 120,000 Australian adults). Panellists, selected at random from the base sample, received an email inviting them to take part in a survey, which included a survey link. Once a panel member clicked on the link and logged in, they were directed to the survey most relevant to them available on the platform at the time, according to the sample definition and quotas based on census data. A plain language statement appeared on screen and respondents were required to electronically consent prior to the survey questions appearing. Proportional quota sampling was used to ensure that respondents were demographically representative of the Victorian adult population, with quotas based on age, gender, household income, location (state and metropolitan or regional) and whether a language other than English is spoken at home.

The study was by approved by the University of Melbourne Human Research Ethics Committee (2056694). Ethics approval applied to all study sites.

## III. RESULTS

Our analysis focused on the urban areas of Sydney and Melbourne during a pre-COVID period (which we use as a baseline), during the first pandemic wave in March and April 2020 (with Australia-wide transmission and mobility restrictions), and during the second wave from July 2020 (with substantial transmission and mobility restrictions in Melbourne but not in Sydney). We identify positive correlations between income security and changes to internet activity during COVID-19. These correlations are consistent with the hypothesis that higher income security is associated with more people working from home during lockdown. This hypothesis is further supported by individual-level data from the CARE survey. We observe that in Sydney this trend persists after the release of lockdown restrictions, indicating the possibility of a ‘new normal’ of remote working conditions, particularly for occupations associated with higher income security. In Melbourne, we find that the role of children conducting their studies online disrupts these correlations due to an inverse relationship between income security and the proportion of families with children.

### A. Employment, income security, and the ability to work from home

Income security is distributed spatially according to distinct patterns, with high values in the central and northeast suburbs of both Sydney and Melbourne (Figures 2a and 2b). The upper 50% income security quantile (Figure 2c) favours managerial and office-based occupations, while the lower 50% quantile (Figure 2d) contains more service staff and other socially-oriented occupations. The frequency distributions of average income security among SA2s in Sydney and Melbourne (respectively) are provided in the Supporting Information Figure S4, which demonstrates that the distributions in the two regions are not significantlydifferent (two sample t-test, *p* = 0.552). High resolution choropleth maps of income security by region can be found in the Supporting Information Figure S6(a).

**FIG. 2.**
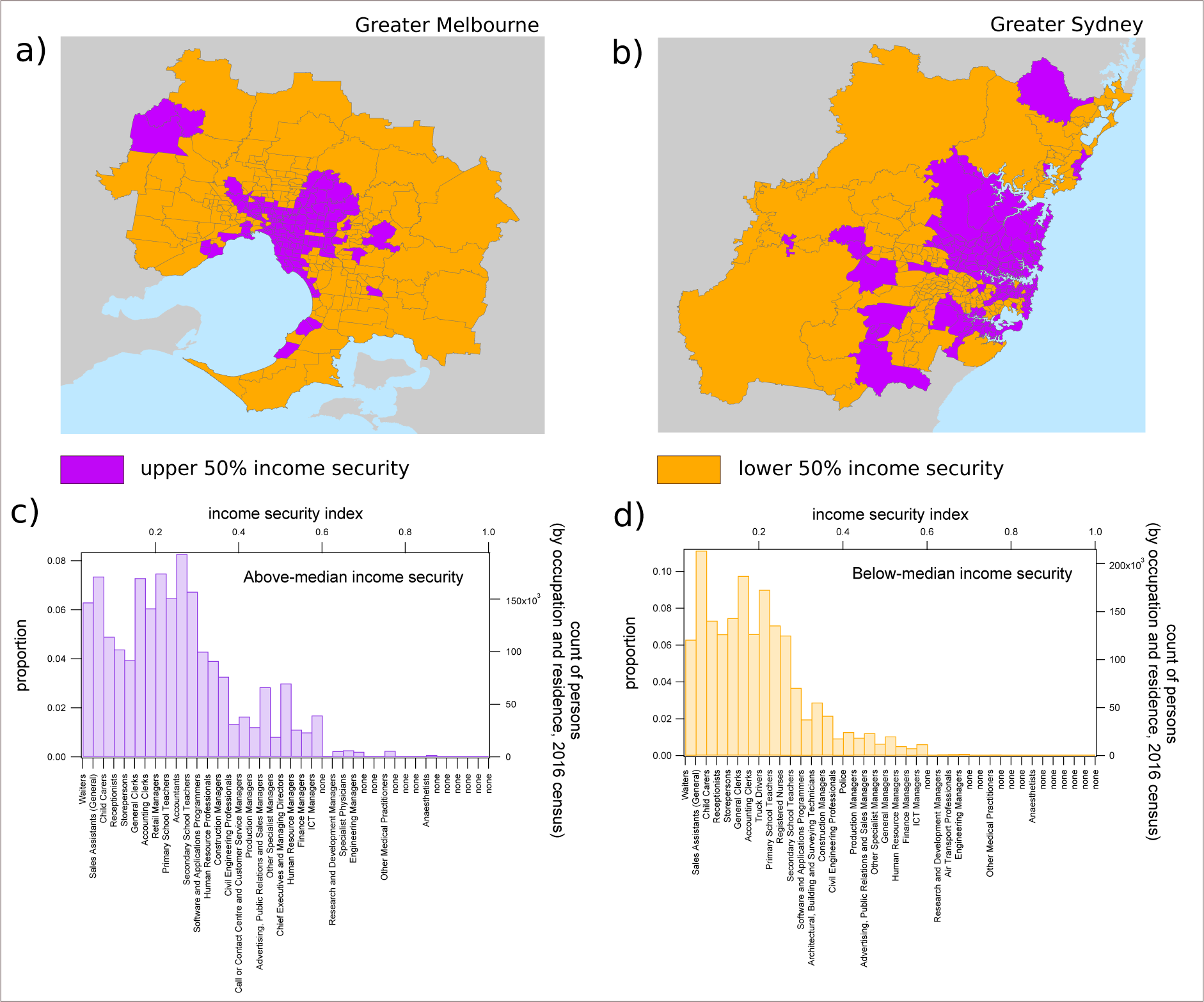
Geospatial and occupational distribution of income security. The choropleth maps in (a) and (b) demonstrate the geospatial distribution of income security in Greater Melbourne and Greater Sydney (respectively) on the scale of Statistical Area Level 2 (SA2). Areas below the median income security index of 0.1809 are colored orange while those above the median are colored purple. The histograms in (c) and (d) demonstrate the distribution of the population in these regions over the income security spectrum, with indicative occupation types for each bin shown in the bottom y-axis labels (the label corresponding to the most prevalent occupation classification for each bin is shown). For both histograms, waiters have the lowest income security, while anaesthetists have the highest.

To examine the qualitative association between income security and the ability to work from home indicated by the distributions in Figures 2c and 2d, we apply the occupation classification method developed by Dingel and Neiman [37]. This results in a binary (0 or 1) value indicating whether or not a particular occupation type can be performed from home. We found a strong association between income security and the ability to work from home (Figure aggregate income security (SA2) (Figure 3b). See the Supporting Information Figure S5 for histograms of the distributions shown in Figure 3a as well as the distributions of the x- and y-axis variables used in Figure 3b.

**FIG. 3.**
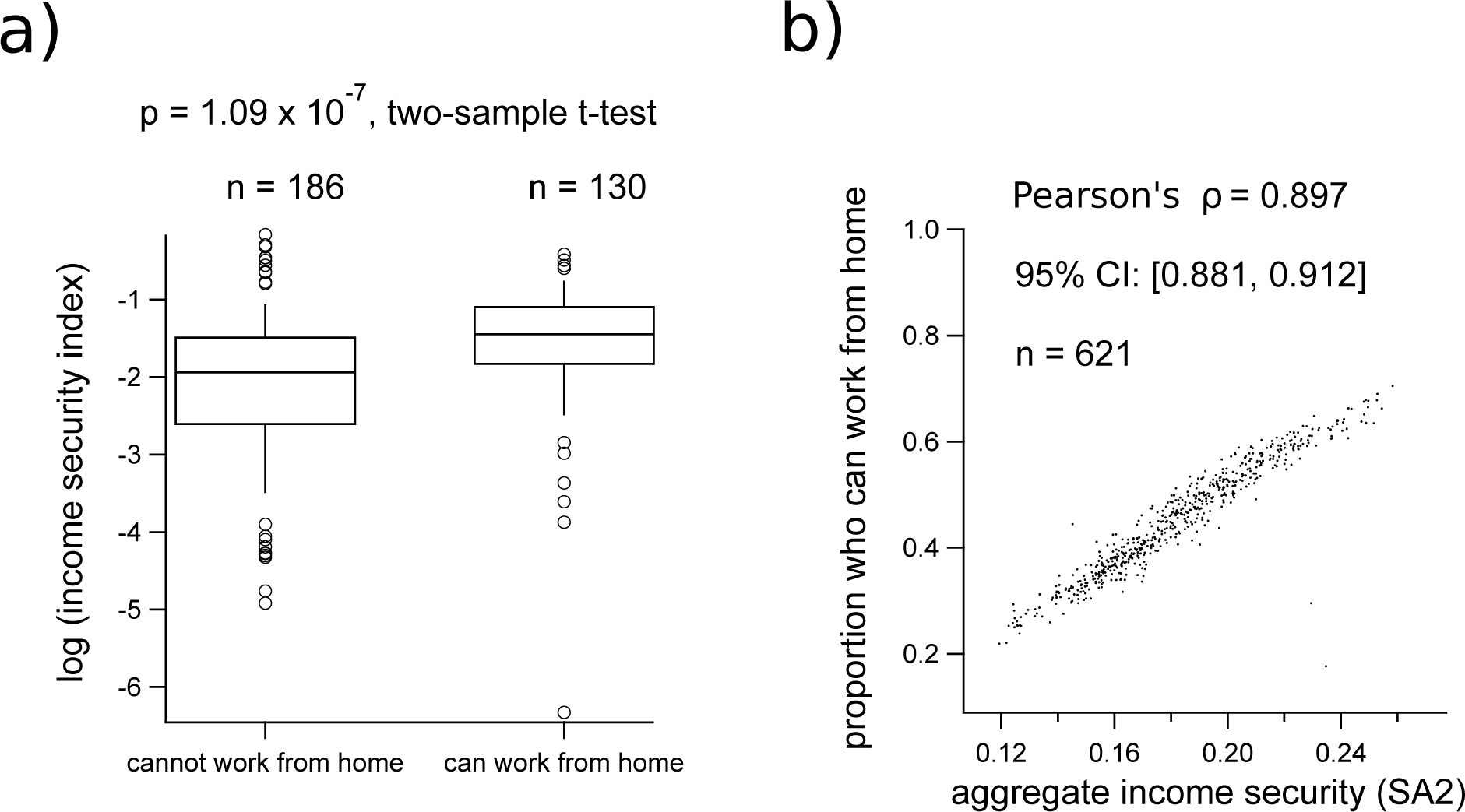
Relationship between income security and the ability to work from home. Box plots in compare the distributions of log-transformed income security, grouped by the ability to work from home for each of 321 occupations classified by 4-digit ANZSCO codes. The distributions in (a) are computed from the HILDA survey, 2018. The scatter plot in (b) demonstrates the correlation between average income security for each SA2 region, and the corresponding average proportion of individuals who can work from home, computed from the occupation distribution of each SA2 in the Greater Sydney and Greater Melbourne regions released in the 2016 ABS Census. [Note: the log-transformed income security data in (a) omits 24 occupations that had a value of 0 for income security (no securely employed HILDA respondents), of these, 2 were grouped into the “can work from home” category and 22 into the “cannot work from home” category. ]

### B. Changes to internet traffic during COVID-19

To quantify changes to home internet use during COVID-19 restrictions, we aggregated internet activity data from all SA2 regions within Greater Sydney and Greater Melbourne (respectively). Over the pre-COVID baseline, we averaged the per-user upload and download rates from the hours of 9am to 12pm in order to capture a baseline measurement of putative remote work-related internet activity (see Methods). During the first and second waves of most restrictive policies, and were followed by increases in total internet use, which peaked approximately one to three weeks after implementation of the tightest level of restrictions (Figure 4).

**FIG. 4.**
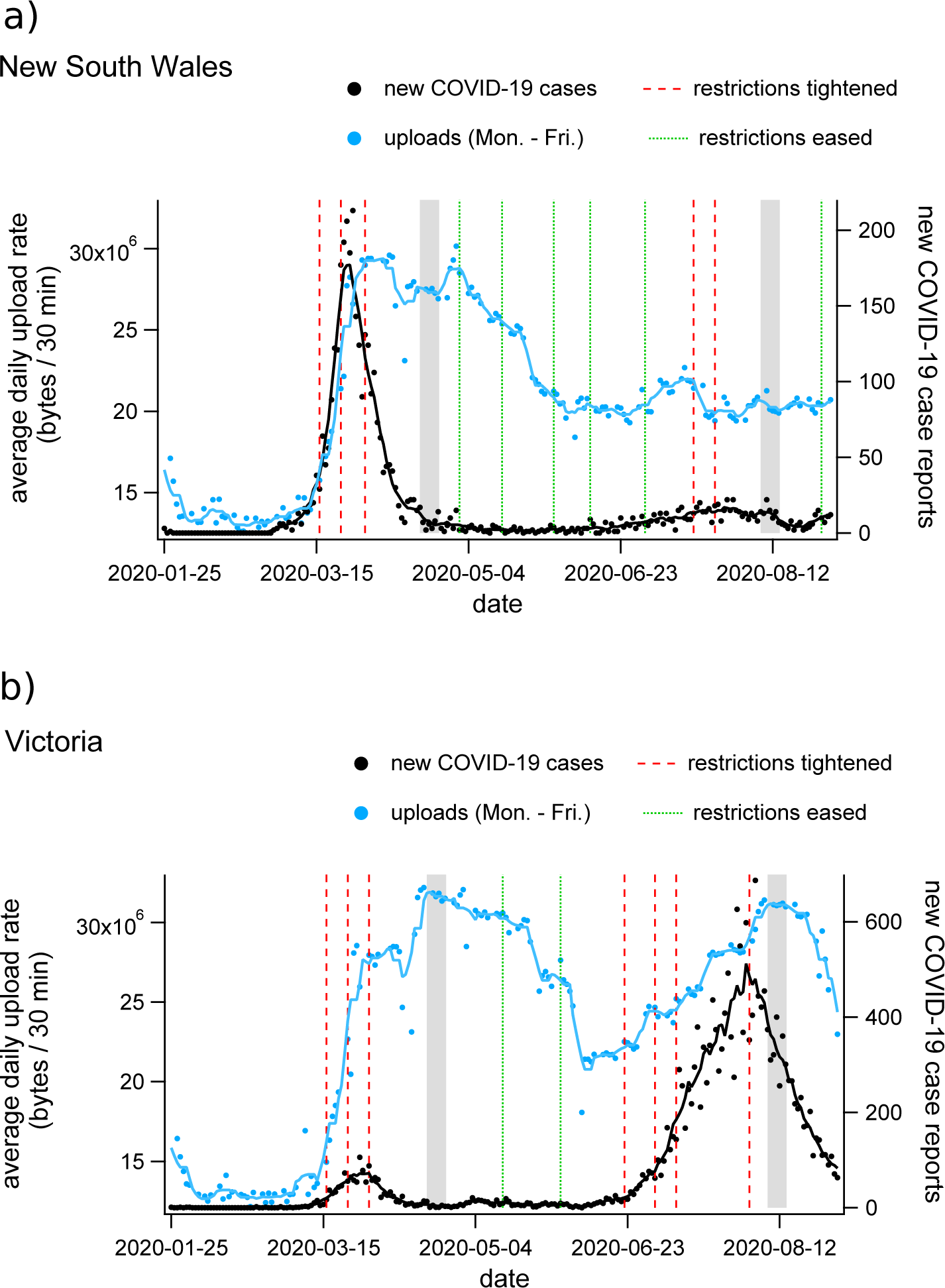
Timeseries plots of average daytime internet use, COVID-19 case incidence, and re- striction policy implementation for (a) New South Wales and (b) Victoria, Australia. Daily average upload rates per household, per 30 min interval between 9am and 12pm are shown as blue dots for weekdays (blue dots, the blue line is the 7-day average). Daily case incidence is shown as black dots (the black line is the 7 day average), and dates on which restriction policies were modified are shown as vertical dashed lines for increasing (red) and decreasing (green) restriction levels. The grey bands indicate the dates over which **nbn**^TM^ data was averaged for our analysis of first- and second-wave changes. See the Supporting Information Figure S1 for an equivalent timeseries presenting average downloads rather than uploads.

After identifying time intervals representative of the changes induced by the first- and second-waves of restrictions, we examined spatial variation among individual SA2 regions during those periods. The grey bands in Figure 4 show the periods over which **nbn**^TM^ data was averaged for each individual SA2 in order to examine the spatial distribution of changes to internet activity during first and second waves of COVID-19. For visualisation of spatial trends, high-resolution choropleth maps of internet activity changes relative to baseline can be found in the Supporting Information Figure S6(b,c).

We found that during the first period of restrictions, areas with higher income security tended to exhibit larger increases in internet volume per household (Figures 5a, 5b, and 5c). However, these trends were produced by qualitatively different changes for downloads and uploads, respectively:

- During the pre-COVID baseline period, absolute download volume tended to decrease with income security, becoming uncorrelated during the first wave of restrictions (Figure 5a). This transition produces larger increases in download volume in areas with higher average income security (Figure 5b, c).
- On the other hand, absolute upload volume shows baseline rates that are initially uncorrelated with income security and transition to an increasing trend during the first wave of restrictions (Figure 5d). This produces changes in upload volume that have similar correlations with income security to those observed for downloads (Figures 5e and 5f), but that occur due to the emergence of a positive correlation rather than the removal of a negative correlation with the onset of restrictions.

The negative baseline trend of download rates with income security may result from the activity of children. We observed a strong positive correlation between the proportion of families with children and baseline download rates (*ρ* = 0.72 95%CI [0.68, 0.75]), and a negative correlation between the proportion of families with children and income security (*ρ* = *−* 0.39 95%CI [*−*0.45*, −* 0.32]). This suggests that children engaged in online activity may establish the negative baseline correlation between download rates and income security (Figure 5a).

Children’s activities also appear to influence the changes observed during lockdown. During the time interval selected to represent the first wave of restrictions (April 18th to April 24th), school holidays were still in effect in Greater Sydney while in Melbourne, children had returned to their studies remotely. Because regions with higher income security tend to have a lower proportion of families with children, remote learning activity weakens the positive association between upload activity and income security produced by adults working from home. Correlations between income security, the proportion of families with children, and internet activity in Sydney and Melbourne (respectively) during the first wave of COVID-19 restrictions are shown in the Supporting Information Tables S2 and S3.

For the second wave of COVID-19 (and associated restrictions), we selected the appropriate time period using internet data from Victoria, where the second epidemic wave was concentrated. In Victoria during the second wave, internet activity peaked during the week of August 8th to August 14th. As for the first wave, this home internet activity peak immediately followed the implementation of the highest level of restrictions (Figure 4b).

Because of the substantially different epidemiological and policy situations in Sydney (New South Wales) and Melbourne (Victoria) during the second-wave period (Figure 4), we examined the relationship between internet traffic, lockdown policy, and income security for each city separately. Comparing the two cities provides insight regarding changes in behaviour related to the contrasting scenarios. During the second wave, Greater Sydney experienced a series of localised outbreaks with minimal social restrictions, while in Melbourne there was a large-scale epidemic with mandatory movement restrictions.

While household internet traffic declines in Sydney during the second wave relative to the first wave, the positive correlation between income security and internet activity relative to baseline remains prominent for both downloads (Figures 6a, 6c), and uploads (Figures 6d, 6f). This is despite the absence of formal stay-at-home orders in the Greater Sydney region at that time (though some restrictions on social gatherings remained in place). The time interval between the first and second waves was long enough to support the assertion that behavioural changes made in response to COVID-19 lockdown policies remain observable after those policies have been formally relaxed.

Greater Melbourne behaves similarly in both waves with respect to changes in download traffic as a function of income security (compare Figure 5(a, c) to Figure 6(a, b)). However, changes to upload volumes do not mirror the correlations observed during the first wave (compare Figure 5(d, f) to Figure 6(d, e)). In fact, there are many areas of Melbourne with high income security that show substantial reductions in upload traffic during the second wave, relative to the first. While our data gives no immediate explanation for this counter-intuitive trend, we speculate that it may be due to alterations in work habits that occurred as the lockdown became protracted. Decreases in upload traffic without corresponding decreases in download traffic could result from individuals continuing to perform work activities from home, but participating in less “face-to-face” online interaction. Conversely, widespread adoption of remote schooling practices could help explain the increase in upload rates for regions in the mid-range of the income security spectrum. Such an effect is consistent with the weak but positive correlations between the proportion of families with children and changes to upload volumes (*ρ* = 0.15 95% CI [0.033, 0.25], see Supporting Information Table S5). This suggestion is also consistent with the observation that daytime internet activity increases during school holidays, when children are more likely to be in the home (Figure 1b).

We hypothesise that schooling in the home had a greater impact on internet volume in general, and upload rates in particular, than working remotely from home during the second wave of COVID-19 restrictions in Melbourne. This hypothesis is supported by a preliminary principal component analysis, summarised in the Supporting Information Figure S2. This 3- component PCA shows an increased role of children in determining upload rates in Greater Melbourne during the second-wave period. Specifically, Figure S2 demonstrates a qualitative change in the relationship between the proportion of families with children, income security, and second-wave changes to upload activity. Upload rate is positively associated with the proportion of families with children in the first component (explaining 55% of the variance) and positively associated with income security in the second component (explaining 32% of the variance). In both components, income security and the proportion of families with children are negatively associated. These PCA results support the suggestion that occupation-related correlations between net changes to upload activity and income security are disrupted by the activities of children in online schooling. This may be explained as a competing effect because lower average income security is associated with a higher proportion of families with children (*ρ* = *−*0.45 95% CI [*−*0.53*, −*0.35], Table S5), while the capacity to work from home increases with income security (*ρ* = 0.90 95% CI [0.88, 0.91], Figure 3b).

### C. Change of work environment for above- and below-median income earners

To confirm that the household-level trends inferred from SA2-level aggregate variables corresponded to observations made on the individual level, we analysed representative data from Victoria collected by the CARE study. While the CARE study did not collect data on income security *per-se*, it did record the annual income bracket reported by each respondent. One of the survey questions was posed as follows: ‘Have you personally experienced a change in work environment (working from home) because of COVID-19 and the measures to CARE survey results for income and working from home due to COVID-19 (Victoria) prevent its spread? (yes or no)’. We computed the proportion of respondents who reported income above and below (or within) the median income bracket for the sample (sample median annual income was $AUD 60,000 to 69,999) who answered ‘yes’ to this question. We then performed a two-tailed Fisher’s exact test to determine the resulting odds ratio between the two groups, and its statistical significance given the response numbers (see Table I). The results demonstrate a strong positive relationship between income and switching to work from home, with an odds ratio of 2.15 (95% CI [1.59, 2.92], *p* = 6.8*×*10*^−^*^7^) computed for the above-median income group, relative to the median-and-below income group. While the income data tabulated by the CARE survey is not an exact representation of the income security score used in our analysis of internet trends (which incorporated contract classification), this result supports the same conclusion: those with higher financial security have more capacity to change their work environments in response to COVID-19 restrictions.

**TABLE I.** From the CARE survey: comparison between above and below-median income groups’ responses to the question: ‘Have you personally experienced a change in work environment (working from home) because of COVID-19 and the measures to prevent its spread? (yes or no)’

| changed work environment? | yes | no | total |
| --- | --- | --- | --- |
| above median income | 144 | 267 | 411 |
| below median income | 93 | 371 | 464 |
| total | 237 | 638 | 875 |

In summary, our results support the hypothesis that occupational factors link the ability to work from home with income security, and clearly show how this link produces strong positive correlations between income security and increases to home internet activity during COVID-19 restrictions. These correlations are consistent with the assertion that higher income security is associated with more people working from home during lockdown. This assertion is further supported by individual-level data from the CARE survey. We observe that in Sydney this trend persists after the release of lockdown restrictions, indicating the possibility of a ‘new normal’ of remote working conditions, particularly for occupations associated with higher income security. In Melbourne, we find that the role of children conducting their studies online disrupts these correlations due to an inverse relationship between income security and the proportion of families with children.

## IV. DISCUSSION

By combining an analysis of occupational factors and distributions with large-scale, high-resolution, real-time data on internet activity, we have broadly characterised the impact of COVID-19 restrictions on two major urban centres in Australia, demonstrating three main findings. First, that occupations associated with greater income security are also associated with the ability to work from home. Second, that Internet usage increased during periods in which COVID-19 restrictions were in place. Increases were greatest in regions with high income security, suggesting that they may be caused by people who were able to adapt to working from home. Finally, that during the second wave in Melbourne, lower-income regions also displayed increased internet usage, likely driven by increased levels of remote schooling.

These findings confirm and elaborate on the general observation that COVID-19 and the associated restrictions on human activity distort normal life activities, with the relative impact largely determined by occupational and demographic factors [39–42]. This unequal impact is dominated by two major influences: the types of occupations in which people are engaged, and the compositions of households and families. Our analysis helps to illustrate how life changed during lockdown. For households with no children, and members engaged work that could be conducted from home, the internet provided a means of continuing livelihood during the prolonged periods of mobility restrictions implemented to combat virus transmission. Furthermore, the members of such housholds were likely to have been previously employed in high-income occupations with relatively strong employment guarantees, adding a measure of confidence in the ability to financially out-last the economic downturn. On the other hand, households with lower income security were also less likely to have been able to work from home, and were more likely to have had children who required care during school closures. For such households, life during lockdown was financially insecure, potentially stressful on the family, and came with a heightened risk of exposure to the pandemic virus due to the work requirements of those occupations which remained active.

### A. Study limitations

Due to the nature of the data we analysed, our study has several limitations. With the exception of the CARE survey results, all of the data analysed in this work is aggregated to sub-populations. Therefore, a direct behavioural interpretation of the correlations we report is contingent on the assumption that the variables we investigate are independently distributed within these sub-populations. While there are likely to be exceptions, the spatial aggregation of areas by income security (Figures 2a and 2b), suggests that the spatial resolution of SA2 regions is sufficient to sample within the boundaries that define salient heterogeneity of the population for the purposes of our study. To confirm this quantitatively, the spatial autocorrelation (Moran’s I) between neighbouring SA2 regions is shown in the Supporting Information section S1 E. This analysis shows highly significant spatial autocorrelation of all relevant variables (income security, and changes to internet usage volumes). From this we conclude that the SA2 scale is an appropriate resolution for the spatially varying quantities studied in this work.

Another inherent limitation is introduced though the use of household internet data in quantifying behaviour across the income spectrum: home internet connections have financial requirements including usage fees and installation costs that may be prohibitive for those at the low end of the income spectrum. For example, a study in the United States recently determined that household internet speeds typically increase with income, and the combination of both high income and high-speed internet is associated with an enhanced ability to self-isolate during the pandemic [21]. Our analysis of relative changes in data volume per active connection addresses potential inequity in the spatial distribution of broadband infrastructure as well as potential correlation of available bandwidth and income security. However, it does not address the possibility that lower-income households use access plans with smaller usage caps. Such a trend could complicate the interpretation of changes to internet volume with respect to working from home during lockdown. The analysis presented here assumes that larger increases to internet usage are indicative of a higher proportion of resident individuals switching to home-based work. An alternative interpretation of the observed trends could be that the proportion of individuals working from home does not depend on income security, but that households with higher income security have access plans with higher usage caps. This alternative interpretation requires data caps to regularly limit internet usage. While such a scenario is possible, we believe it to be less plausible than the one we chose. In addition, the individual-level CARE survey analysis supports our chosen interpretation.

A related limitation is that households with no internet connection were implicitly excluded from our analysis. Using Australian data from the 2016 ABS Census, we computed a correlation of *ρ* = 0.50, 95% CI [0.44, 0.56], associating the fraction of households with an internet connection (as of 2016) with the income security measure computed here (see Supporting Information Table S1). Therefore, our use of home internet traffic data to estimate the ability of workers with differing income security to adapt to COVID-19 restrictions may omit the behaviour of many low-income households, producing an underestimate the effects of occupational factors on behaviour during the crisis.

## Data Availability

All processed data necessary for reproducing the figures and results of this paper is available in the GitHub repository located here: https://github.com/cjzachreson/Internet_Income_and_COVID19_in_Australia. Access to raw data from the HILDA survey, nbn(TM), and CARE survey was obtained under restricted access agreements and cannot be made directly available. However, arrangements for access may be made for eligible researchers. All ABS data can be accessed by following the instructions provided here: https://www.abs.gov.au/websitedbs/D3310114.nsf/home/How+to+Apply+for+Microdata.

https://www.abs.gov.au/websitedbs/D3310114.nsf/home/How+to+Apply+for+Microdata

## V. ACKNOWLEDGEMENTS

CZ, NG, and MT were supported by an internal seed grant from the Melbourne School of Engineering, of the University of Melbourne. We gratefully acknowledge the contribution of **nbn** co ltd. who provided access to the internet usage data, and in particular the data expertise and extraction of Michael Joyce, Jason Godden and Neil Sequeira. This publication uses data collected as part of the COVID-19 Attitudes Resilience and Epidemiology (CARE) Study. We would like to acknowledge all of the study participants, and the University of Melbourne and Centre for Ethnicity, Culture and Health staff who helped to make this study possible. We acknowledge the work of the investigator team: Lisa Gibbs, David Price, Katitza Marinkovic Chavez, Niamh Meagher, Lauren Carpenter and Colin MacDougall. The CARE Study was funded by the Melbourne School of Population and Global Health and the Optimise Study. We thank Tara Purcell for assembling information on Australian national and state COVID- 19 response measures.This publication uses data collected from The Household, Income and Labour Dynamics in Australia (HILDA) Survey. We would like to acknowledge all of the participants and The Melbourne Institute, The University of Melbourne who made this survey possible.

## VI DATA AVAILABILITY

All processed data necessary for reproducing the figures and results of this paper is available in the GitHub repository located here: https://github.com/cjzachreson/Internet_ Income_and_COVID19_in_Australia. Access to raw data from the HILDA survey, **nbn**^TM^, and CARE survey was obtained under restricted access agreements and cannot be made directly available. However, arrangements for access may be made for eligible researchers. All ABS data can be accessed by following the instructions provided here: https://www.abs.gov.au/websitedbs/D3310114.nsf/home/How+to+Apply+for+Microdata.

## VII AUTHOR CONTRIBUTIONS

All authors contributed to study design, data interpretation, and manuscript preparation. CZ composed the manuscript and figures, and performed all statistical analysis. MT performed pre-processing of **nbn**^TM^ data. EM performed pre-processing of HILDA survey data and computed work-from-home classifications for each occupation. FMS provided CARE survey data and assisted in its interpretation. NG, MT, and RB conceived of and initialised the study.

## S1. SUPPORTING INFORMATION

### A. Timeseries of case incidence and download rates

**FIG. S1.**
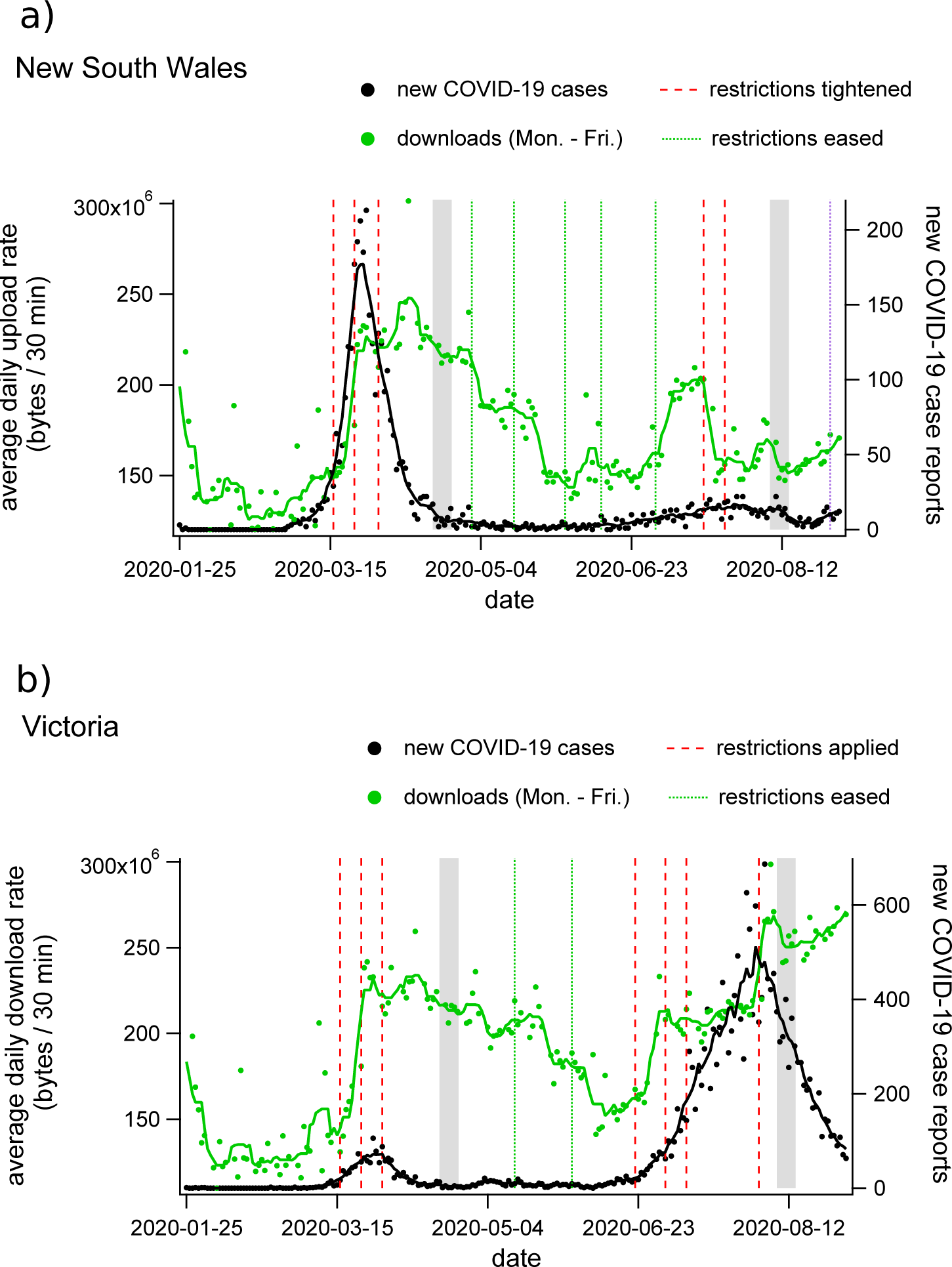
Timeseries plots of average daytime internet use, COVID-19 case incidence, and restriction policy implementation for (a) New South Wales and (b) Victoria. Daily average download rates per household, per 30 min interval between 9am and 12pm are shown as blue dots for weekdays (green dots, the green line is the 7-day average). Daily case incidence is shown as black dots (the black line is the 7 day average), and dates on which restriction policies were modified are shown as vertical dashed lines for increasing (red) and decreasing (green) restriction levels. The grey bands indicate the dates over which **nbn**^TM^ data was averaged for our analysis of 1st- and 2nd-wave changes.

### B. Correlations with other demographic factors

Tables S1 through S5 show the correlation (Pearson’s *ρ*, *±*95% CI bounds) between various measures of internet traffic and several alternate demographic factors that we considered in our study in addition to income security.

These are:

- *p_W FH_* (*MT W P* ), the proportion of individuals from each region who reported working from home in the 2016 Australian Census counts of Method of Travel to Work (MTWP). We selected this measure because it serves as an alternate measurement of the tendency to work from home under normal circumstances and could plausibly explain baseline internet traffic.
- *p_internet_*, the proportion of households with an internet connection, as counted by the 2016 Australian Census. We chose this measure because it provides an alternate indicator of the importance of internet in normal activities.
- *p_children_*, the proportion of families with children, as determined by the 2016 Australian Census. We chose this measure for two reasons. Firstly, because baseline internet traffic could plausibly depend on the size of a family and the presence of children and secondly, because it may help elucidate the effects of school closures on COVID-19 usage levels.

Tables S1 through S5 demonstrate the correlations of internet traffic measures with each of these factors, as well as the correlations between the demographic factors. Table S1 shows correlations between factors and internet usage for SA2 regions in Greater Sydney and Greater Melbourne during the first wave of COVID-19 restrictions. Notably, the *p_children_* factor appears to explain the negative correlation between baseline downloads and income security. Income security is negatively correlated with the proportion of families with children, while baseline download traffic is positively correlated. This observation indicates that the presence of children may increase internet use under normal circumstances.

The case is different for baseline upload traffic, where correlation is negligible with both income security and *p_children_*. This distinction between upload and download traffic in the baseline data suggests that typical internet activities undertaken by children do not involve significant amounts of out-bound streaming, or two-way communication. Table S1 also shows that while download traffic during the first wave of COVID-19 restrictions correlates with the proportion of children, the change in download traffic relative to baseline is negatively correlated. On the other hand, changes in download traffic relative to baseline correlate strongly with income security indicating that occupational factors associated with working from home dominate over the contributions of children with respect to relative changes in download activity due to COVID-19 lockdown measures. This contrast is even more apparent when examining upload traffic, for which factors associated with working from home dominate all measures of COVID-19 related changes in traffic.

Tables S2 and S3 show these correlations for Greater Sydney and Greater Melbourne, respectively, and indicate that while the two regions have qualitatively similar correlations between changes in upload volume and income security, Melbourne demonstrates a much less pronounced correspondence. We speculate that this is due to the activities of school children. During this period, students in Greater Sydney were not attending class due to the mid-semester break, while in Melbourne school was in session but schools buildings were closed and most classes were conducted remotely. The negative correlation between income security and the proportion of families with children suggests that increases in home-learning activity could disrupt occupation-related correlations between income security and increased internet traffic.

Tables S4 and S5 show correlations between demographic factors and internet traffic in the Greater Sydney and Greater Melbourne regions, respectively, during the 2nd wave of COVID- 19 restrictions. During this wave, Greater Sydney did not have large numbers of cases and did not go through a second wave of mandated social distancing measures. Importantly, almost all schools were open during this period, with a few exceptions where localised outbreaks were detected in students. Comparison of correlations between internet use, income security, and *p_children_* indicate that work activities were the primary driver of above-baseline internet traffic during this period. The difference is particularly stark when examining upload behaviour, which is strongly positively correlated to income security and weakly negatively correlated to *p_children_*, which suggests that deviations from baseline uploads may be a strong indicator for work-from-home behaviour, at least when schools are open.

On the other hand, Greater Melbourne shows unique correlations during the 2nd wave of COVID-19 restrictions. In Melbourne, classes in primary and secondary education were held remotely during the 1st and 2nd waves of COVID-19 restrictions, and positive correlation between income security and increased upload rates is not observed. In this case, the activity of children appears to dominate changes to internet traffic, nullifying the occupation-related correlation between income security and changes to upload rates.

**TABLE S1.** Corrleation (Pearson’s *ρ*) of internet usage with demographic variables (Greater Sydney, COVID-19 1st wave)

| | income security | $p_{WFH}(MTWP)$ | $p_{internet}$ | $p_{children}$ |
| --- | --- | --- | --- | --- |
| $ds_{baseline}$ | -0.49 [-0.55, -0.43] | -0.51 [-0.57, -0.45] | 0.062 [-0.019, 0.14] | 0.72 [0.68, 0.75] |
| $ds_{peak1}$ | 0.031 [-0.05, 0.11] | -0.088 [-0.17, -0.0079] | 0.52 [0.46, 0.58] | 0.69 [0.64, 0.73] |
| $\Delta ds_{peak1}$ | 0.59 [0.53, 0.64] | 0.45 [0.38, 0.51] | 0.71 [0.66, 0.75] | 0.2 [0.12, 0.27] |
| $\Delta ds_{peak1}/ds_{baseline}$ | 0.77 [0.73, 0.8] | 0.66 [0.62, 0.71] | 0.49 [0.42, 0.55] | -0.27 [-0.34, -0.19] |
| $us_{baseline}$ | 0.12 [0.042, 0.2] | 0.082 [0.0011, 0.16] | 0.19 [0.11, 0.26] | 0.13 [0.048, 0.21] |
| $us_{peak1}$ | 0.58 [0.52, 0.63] | 0.38 [0.31, 0.45] | 0.49 [0.42, 0.55] | -0.082 [-0.16, -0.0013] |
| $\Delta us_{peak1}$ | 0.54 [0.48, 0.59] | 0.39 [0.32, 0.45] | 0.44 [0.37, 0.5] | -0.11 [-0.19, -0.035] |
| $\Delta us_{peak1}/us_{baseline}$ | 0.37 [0.3, 0.44] | 0.26 [0.19, 0.34] | 0.25 [0.17, 0.32] | -0.17 [-0.25, -0.094] |
| income security | 1 | 0.7 [0.65, 0.74] | 0.5 [0.44, 0.56] | -0.39 [-0.45, -0.32] |
| $p_{WFH}(MTWP)$ | 0.7 [0.65, 0.74] | 1 | 0.43 [0.36, 0.49] | -0.31 [-0.38, -0.23] |
| $p_{internet}$ | 0.5 [0.44, 0.56] | 0.43 [0.36, 0.49] | 1 | 0.33 [0.26, 0.4] |
| $p_{Children}$ | -0.39 [-0.45, -0.32] | -0.31 [-0.38, -0.23] | 0.33 [0.26, 0.4] | 1 |

**TABLE S2.** Corrleation (Pearson’s *ρ*) of internet usage with demographic variables (Greater Melbourne, COVID-19 1st wave)

| | income security | $p_{WFH}(MTWP)$ | $p_{internet}$ | $p_{children}$ |
| --- | --- | --- | --- | --- |
| $ds_{baseline}$ | -0.57 [-0.64, -0.49] | -0.57 [-0.64, -0.49] | -0.081 [-0.19, 0.033] | 0.72 [0.66, 0.77] |
| $ds_{peak1}$ | 0.027 [-0.088, 0.14] | -0.12 [-0.24, -0.01] | 0.42 [0.32, 0.51] | 0.69 [0.62, 0.74] |
| $\Delta ds_{peak1}$ | 0.63 [0.56, 0.7] | 0.38 [0.28, 0.47] | 0.72 [0.66, 0.77] | 0.17 [0.059, 0.28] |
| $\Delta ds_{peak1}/ds_{baseline}$ | 0.79 [0.74, 0.83] | 0.61 [0.54, 0.68] | 0.53 [0.44, 0.61] | -0.23 [-0.33, -0.11] |
| $us_{baseline}$ | 0.18 [0.064, 0.29] | 0.12 [0.0048, 0.23] | 0.17 [0.057, 0.28] | 0.012 [-0.1, 0.13] |
| $us_{peak1}$ | 0.81 [0.76, 0.84] | 0.5 [0.41, 0.58] | 0.55 [0.46, 0.62] | -0.18 [-0.29, -0.066] |
| $\Delta us_{peak1}$ | 0.78 [0.73, 0.82] | 0.47 [0.37, 0.55] | 0.53 [0.44, 0.6] | -0.16 [-0.27, -0.05] |
| $\Delta us_{peak1}/us_{baseline}$ | 0.64 [0.57, 0.7] | 0.37 [0.27, 0.47] | 0.39 [0.29, 0.48] | -0.19 [-0.3, -0.074] |
| income security | 1 | 0.65 [0.58, 0.72] | 0.58 [0.5, 0.65] | -0.36 [-0.45, -0.25] |
| $p_{WFH}(MTWP)$ | 0.65 [0.58, 0.72] | 1 | 0.41 [0.31, 0.5] | -0.31 [-0.41, -0.2] |
| $p_{internet}$ | 0.58 [0.5, 0.65] | 0.41 [0.31, 0.5] | 1 | 0.27 [0.16, 0.38] |
| $p_{Children}$ | -0.36 [-0.45, -0.25] | -0.31 [-0.41, -0.2] | 0.27 [0.16, 0.38] | 1 |

**TABLE S3.** Corrleation (Pearson’s *ρ*) of internet usage with demographic variables
(Greater Melbourne, COVID-19 1st wave)

| | income security | $p_{WFH}(MTWP)$ | $p_{internet}$ | $p_{children}$ |
| --- | --- | --- | --- | --- |
| $ds_{baseline}$ | -0.46 [-0.55, -0.37] | -0.47 [-0.55, -0.37] | 0.19 [0.08, 0.3] | 0.71 [0.65, 0.76] |
| $ds_{peak1}$ | 0.015 [-0.098, 0.13] | -0.071 [-0.18, 0.043] | 0.61 [0.54, 0.68] | 0.69 [0.62, 0.74] |
| $\Delta ds_{peak1}$ | 0.55 [0.46, 0.62] | 0.48 [0.39, 0.56] | 0.7 [0.63, 0.75] | 0.24 [0.13, 0.34] |
| $\Delta ds_{peak1}/ds_{baseline}$ | 0.76 [0.71, 0.8] | 0.72 [0.66, 0.77] | 0.44 [0.34, 0.53] | -0.3 [-0.4, -0.19] |
| $us_{baseline}$ | 0.04 [-0.074, 0.15] | 0.062 [-0.052, 0.17] | 0.21 [0.099, 0.32] | 0.18 [0.068, 0.29] |
| $us_{peak1}$ | 0.42 [0.32, 0.51] | 0.32 [0.21, 0.42] | 0.44 [0.35, 0.53] | 0.058 [-0.055, 0.17] |
| $\Delta us_{peak1}$ | 0.4 [0.31, 0.49] | 0.31 [0.2, 0.41] | 0.38 [0.28, 0.48] | 0.0016 [-0.11, 0.11] |
| $\Delta us_{peak1}/us_{baseline}$ | 0.26 [0.15, 0.36] | 0.17 [0.057, 0.28] | 0.14 [0.029, 0.25] | -0.1 [-0.21, 0.01] |
| income security | 1 | 0.72 [0.66, 0.77] | 0.42 [0.32, 0.51] | -0.45 [-0.53, -0.35] |
| $p_{WFH}(MTWP)$ | 0.72 [0.66, 0.77] | 1 [1, 1] | 0.41 [0.31, 0.5] | -0.34 [-0.44, -0.24] |
| $p_{internet}$ | 0.42 [0.32, 0.51] | 0.41 [0.31, 0.5] | 1 | 0.39 [0.29, 0.49] |
| $p_{Children}$ | -0.45 [-0.53, -0.35] | -0.34 [-0.44, -0.24] | 0.39 [0.29, 0.49] | 1 |

**TABLE S4.** Correlation (Pearson’s *ρ*) of internet usage with demographic variables (Greater Melbourne, COVID-19 2nd wave)

| | income security | $p_{WFH}(MTWP)$ | $p_{internet}$ | $p_{children}$ |
| --- | --- | --- | --- | --- |
| $ds_{baseline}$ | -0.57 [-0.64, -0.49] | -0.57 [-0.64, -0.49] | -0.081 [-0.19, 0.033] | 0.72 [0.66, 0.77] |
| $ds_{peak2}$ | -0.085 [-0.2, 0.03] | -0.27 [-0.37, -0.16] | 0.33 [0.23, 0.43] | 0.71 [0.65, 0.76] |
| $\Delta ds_{peak2}$ | 0.76 [0.71, 0.8] | 0.42 [0.32, 0.51] | 0.66 [0.59, 0.72] | -0.044 [-0.16, 0.071] |
| $\Delta ds_{peak2}/ds_{baseline}$ | 0.83 [0.79, 0.86] | 0.55 [0.47, 0.63] | 0.45 [0.36, 0.54] | -0.32 [-0.42, -0.21] |
| $us_{baseline}$ | 0.18 [0.064, 0.29] | 0.12 [0.0048, 0.23] | 0.17 [0.057, 0.28] | 0.012 [-0.1, 0.13] |
| $us_{peak2}$ | 0.85 [0.81, 0.88] | 0.47 [0.37, 0.55] | 0.52 [0.43, 0.6] | -0.24 [-0.35, -0.13] |
| $\Delta us_{peak2}$ | 0.84 [0.8, 0.87] | 0.44 [0.35, 0.53] | 0.51 [0.42, 0.59] | -0.23 [-0.34, -0.12] |
| $\Delta us_{peak2}/us_{baseline}$ | 0.74 [0.68, 0.79] | 0.38 [0.28, 0.47] | 0.42 [0.32, 0.51] | -0.24 [-0.35, -0.13] |
| income security | 1 | 0.65 [0.58, 0.72] | 0.58 [0.5, 0.65] | -0.36 [-0.45, -0.25] |
| $p_{WFH}(MTWP)$ | 0.65 [0.58, 0.72] | 1 | 0.41 [0.31, 0.5] | -0.31 [-0.41, -0.2] |
| $p_{internet}$ | 0.58 [0.5, 0.65] | 0.41 [0.31, 0.5] | 1 | 0.27 [0.16, 0.38] |
| $p_{Children}$ | -0.36 [-0.45, -0.25] | -0.31 [-0.41, -0.2] | 0.27 [0.16, 0.38] | 1 |

**TABLE S5.** Correlation (Pearson’s *ρ*) of internet usage with demographic variables (Greater Melbourne, COVID-19 2nd wave)

| | income security | $p_{WFH}(MTWP)$ | $p_{internet}$ | $p_{children}$ |
| --- | --- | --- | --- | --- |
| $ds_{baseline}$ | -0.46 [-0.55, -0.37] | -0.47 [-0.55, -0.37] | 0.19 [0.08, 0.3] | 0.71 [0.65, 0.76] |
| $ds_{peak2}$ | 0.0055 [-0.11, 0.12] | -0.0058 [-0.12, 0.11] | 0.65 [0.58, 0.71] | 0.75 [0.69, 0.79] |
| $\Delta ds_{peak2}$ | 0.35 [0.24, 0.44] | 0.38 [0.27, 0.47] | 0.74 [0.68, 0.79] | 0.49 [0.4, 0.57] |
| $\Delta ds_{peak2}/ds_{baseline}$ | 0.65 [0.58, 0.71] | 0.7 [0.64, 0.75] | 0.51 [0.42, 0.59] | -0.091 [-0.2, 0.023] |
| $us_{baseline}$ | 0.04 [-0.074, 0.15] | 0.062 [-0.052, 0.17] | 0.21 [0.099, 0.32] | 0.18 [0.068, 0.29] |
| $us_{peak2}$ | -0.1 [-0.22, 0.0081] | -0.11 [-0.22, -0.00082] | 0.19 [0.074, 0.29] | 0.21 [0.099, 0.32] |
| $\Delta us_{peak2}$ | -0.11 [-0.22, 0.0032] | -0.12 [-0.23, -0.007] | 0.12 [0.0072, 0.23] | 0.15 [0.033, 0.25] |
| $\Delta us_{peak2}/us_{baseline}$ | -0.068 [-0.18, 0.046] | -0.075 [-0.19, 0.039] | -0.007 [-0.12, 0.11] | 0.018 [-0.095, 0.13] |
| income security | 1 | 0.72 [0.66, 0.77] | 0.42 [0.32, 0.51] | -0.45 [-0.53, -0.35] |
| $p_{WFH}(MTWP)$ | 0.72 [0.66, 0.77] | 1 | 0.41 [0.31, 0.5] | -0.34 [-0.44, -0.24] |
| $p_{internet}$ | 0.42 [0.32, 0.51] | 0.41 [0.31, 0.5] | 1 | 0.39 [0.29, 0.49] |
| $p_{children}$ | -0.45 [-0.53, -0.35] | -0.34 [-0.44, -0.24] | 0.39 [0.29, 0.49] | 1 |

**FIG. S2.**
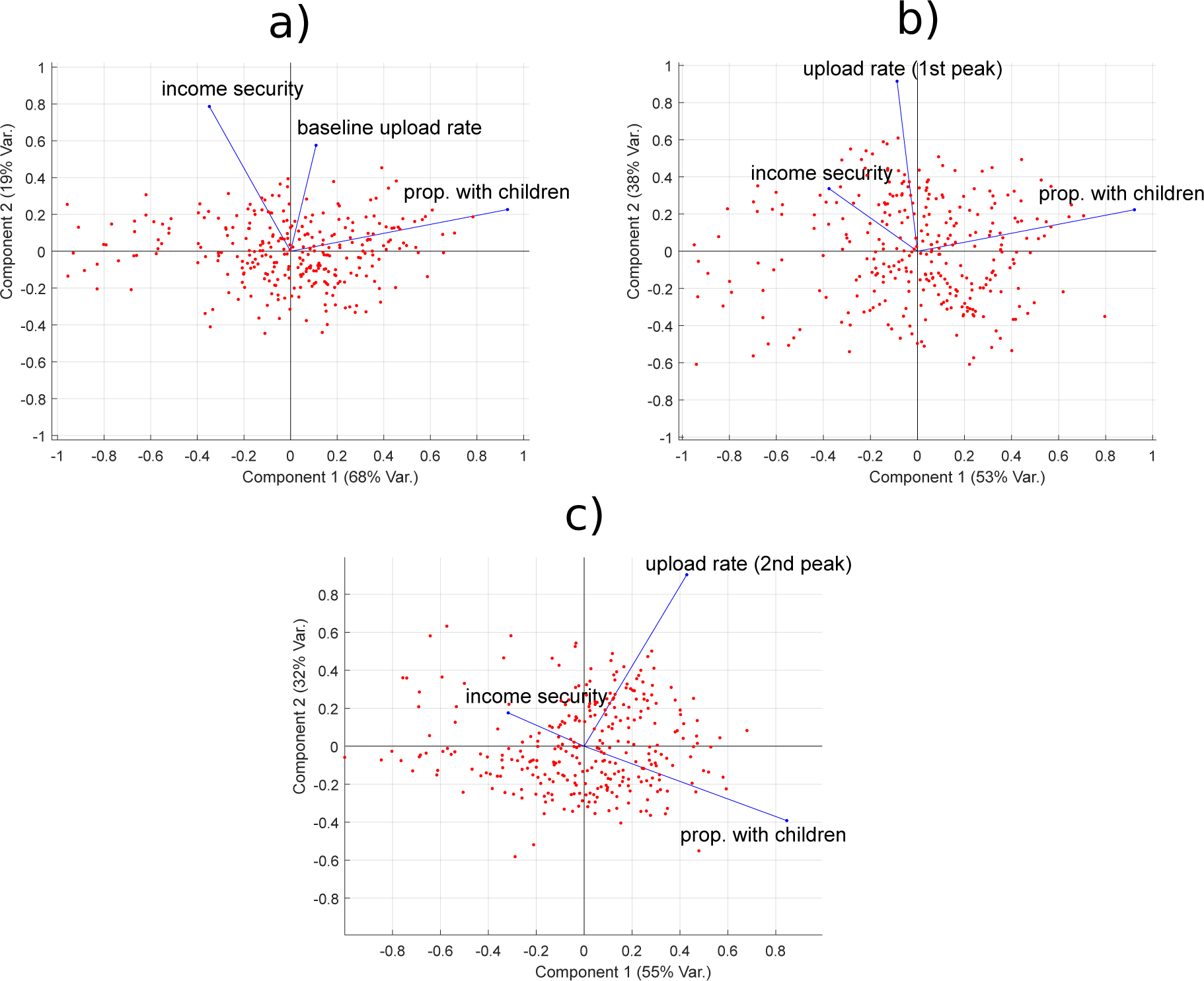
Biplots demonstrating the results of a 3-component principal component analysis for the SA2 regions of Greater Melbourne, using income security, proportion of families with children, and (a) baseline uploads, (b) absolute upload rates during the first wave of COVID- 19 restrictions, and (c) absolute upload rates during the 2nd wave of COVID-19 restrictions. The red dots represent SA2 regions, which are positioned based on the corresponding values of the first two principal components. The blue vector lines represent the contributions of each variable (labeled) to these two components.

### C. Distributions of income security and work-from-home classification by occupation and SA2 region

**FIG. S3.**
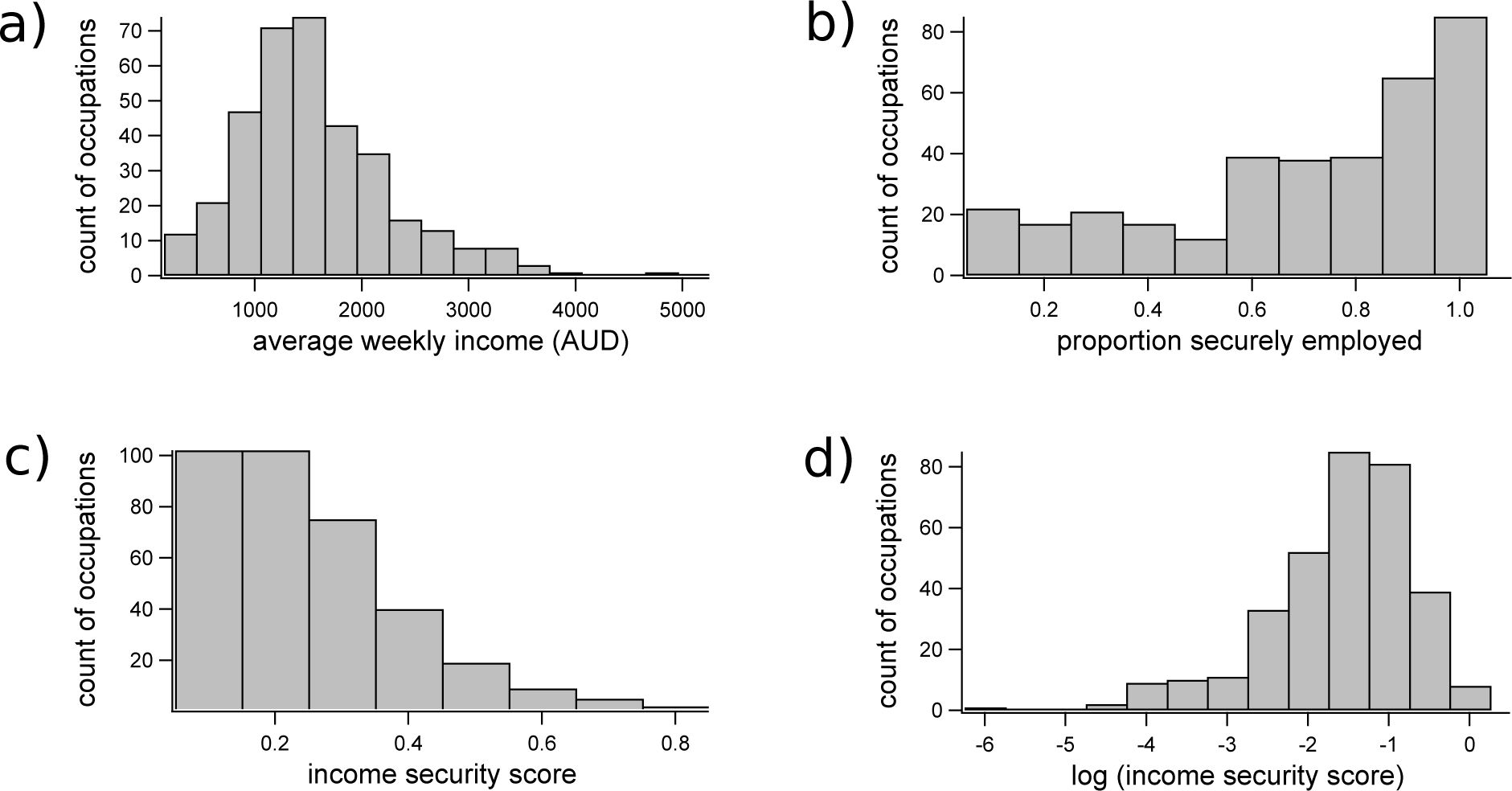
Histograms demonstrating the distributions among occupation classifications of (a) income, (b) proportion securely employed, (c) income security scores computed as the product of relative income and proportion securely employed, and (d) log-transformed income security scores.

**FIG. S4.**
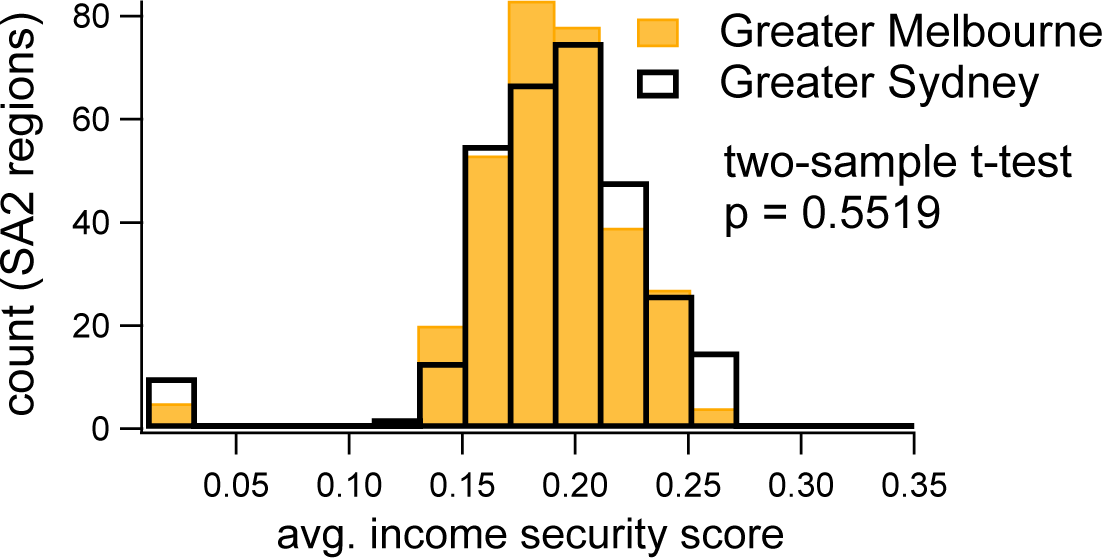
Histograms demonstrating the distribution of average income security scores over the SA2 regions in Greater Melbourne (solid yellow bars) and Greater Sydney (open black bars).

**FIG. S5.**
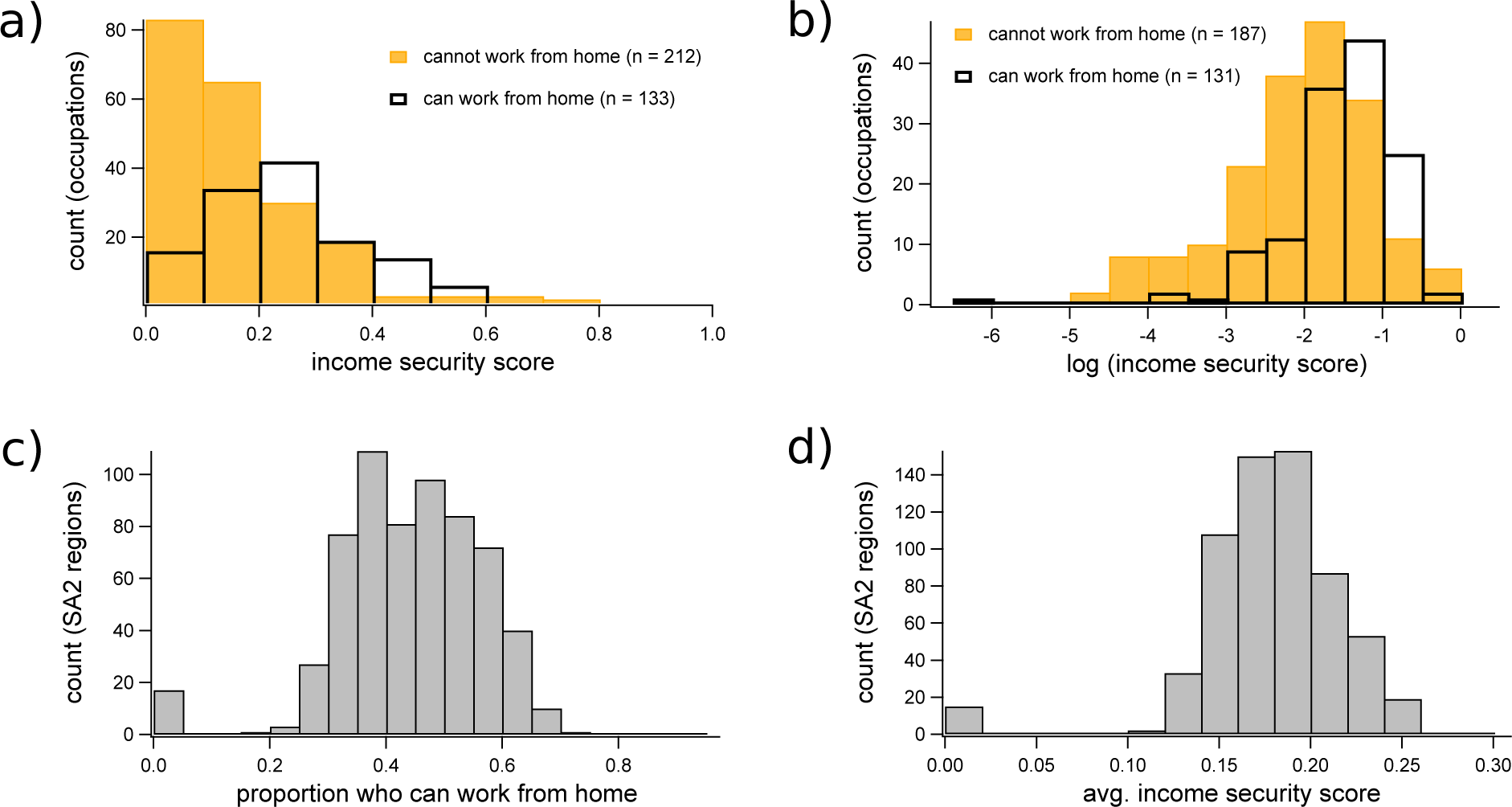
(a, b) Histograms demonstrating the distribution of (a) income security by occupation, grouped by the ability to work from home, with the log-transformed distributions shown in (b). In (a) and (b), open black bars represent occupations for which at least 50% of HILDA respondents were securely employed, while yellow bars represent those occupations for which less than 50% of HILDA respondents were securely employed. (c, d) Histograms showing the distribution among SA2 regions of (c) the proportion of occupied individuals who can work from home, and (d) average income security. The histograms in (c) and (d) include all SA2 regions in Greater Sydney and Greater Melbourne.

### D. Geospatial distribution of income security and changes to internet activity

**FIG. S6.**
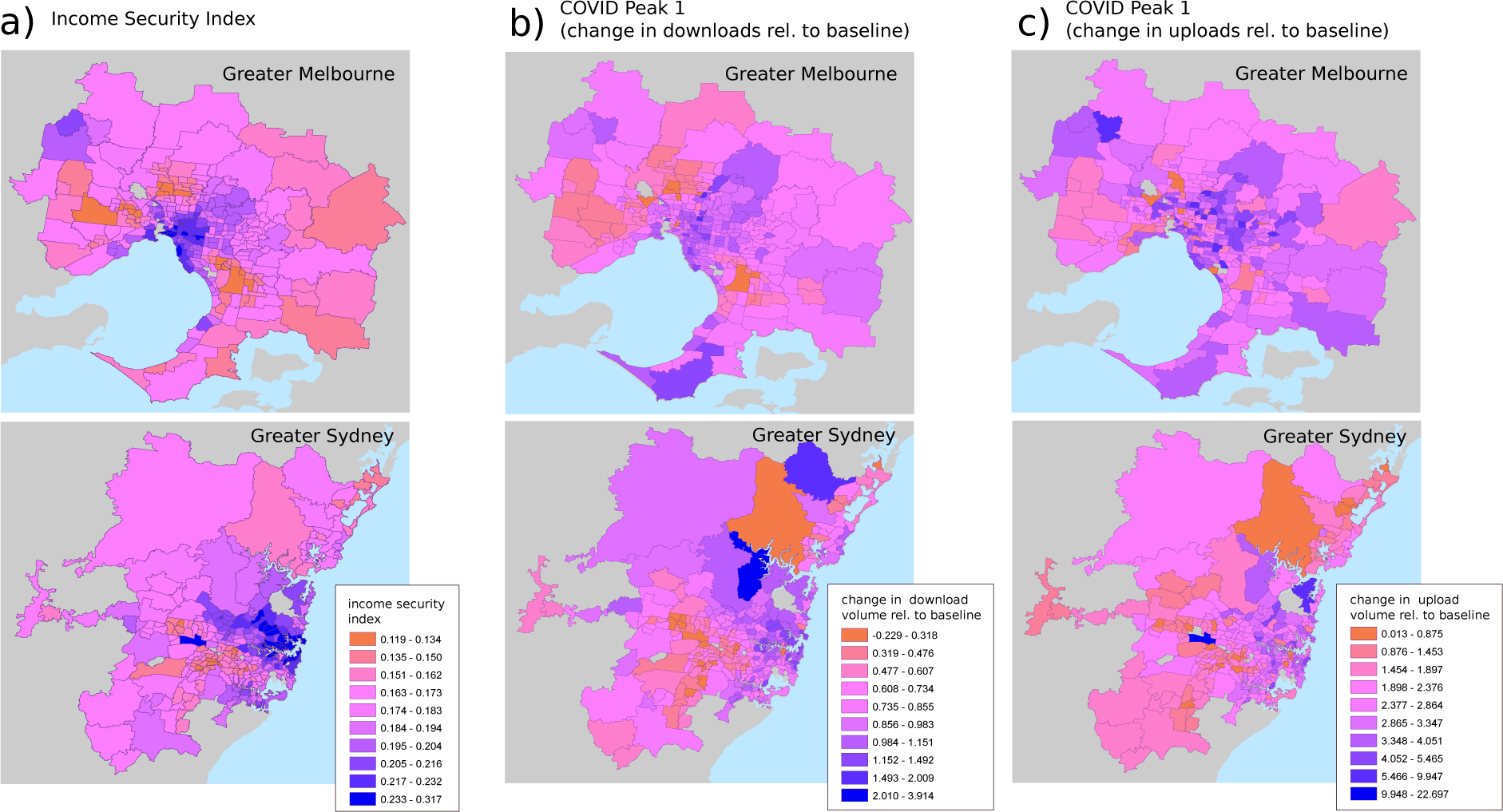
Choropleth maps demonstrating the geospatial distributions of income security and changes to internet activity during the first wave of COVID-19 restrictions in Greater Sydney and Greater Melbourne. The maps in (a) show the spatial distribution of aggregate income security, while (b) and (c) show the spatial distribution of changes in download and upload volumes (respectively) during the first-wave period (April 18th to April 24th, 2020).

### E. Tests of Geospatial Autocorrelation of income security and changes to internet activity

We have computed the Global Moran’s I statistics of income security by SA2, as well as relative changes to internet use at Peak 1 and Peak 2 in both Melbourne and Sydney.

The Moran’s I statistic is a descriptor of the extent to which a variable is spatially auto- correlated, or, conversely, whether the phenomenon studied is randomly distributed in space (which is typically the null hypothesis).

The Moran’s I statistics for both Melbourne and Sydney are shown in Table S6. All statistics have been computed with the assumption of a Queen neighbourhood, with a row- standardised weight distribution between neighbours, using the permutation-based Moran’s I statistic implementation in the spdep package [43, 44] of the R statistical environment, based on 1000 Monte-Carlo runs.

**TABLE S6.** caption

| City | Variable | Moran's I | p-value |
| --- | --- | --- | --- |
| Melbourne | Income security | 0.736 | < .001 |
|  | Peak 1 (US) | 0.251 | < .001 |
|  | Peak 2 (US) | 0.131 | < .001 |
|  | Peak 1 (DS) | 0.574 | < .001 |
|  | Peak 2 (DS) | 0.501 | < .001 |
| Sydney | Income security | 0.786 | < .001 |
|  | Peak 1 (US) | 0.510 | < .001 |
|  | Peak 2 (US) | 0.656 | < .001 |
|  | Peak 1 (DS) | 0.596 | < .001 |
|  | Peak 2 (DS) | 0.588 | < .001 |

Changes to internet use relative to baseline show higher levels of autocorrelation for downloads than for uploads at both peaks in Melbourne (Figure S7(b,c,d,e)), while in Sydney these correlations remain the same (Figure S9(b,c,d,e)). Yet, the permutation-based Moran’s tests confirm that all systems are significantly autocorrelated, as illustrated by the permutation- based density plots in Figures S8 and S10).

**FIG. S7.**
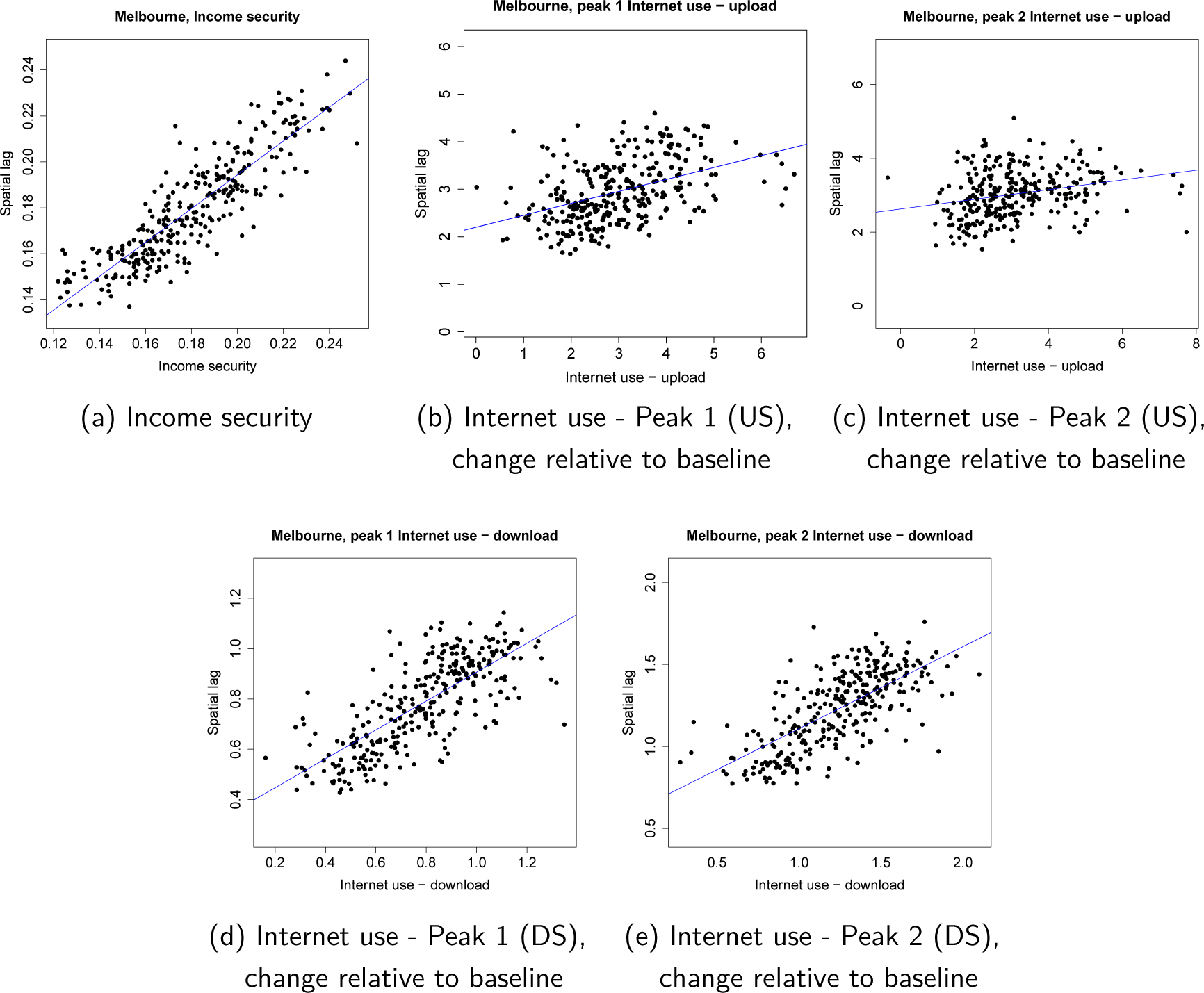
Melbourne - Plots of the variables and their spatially lagged values for income security (a), as well as relative change to upload volume at peak 1 (b), relative change to upload volume at peak 2 (c), relative change to download volume at peak 1 (d), and relative change to download volume at peak 2 (e).

**FIG. S8.**
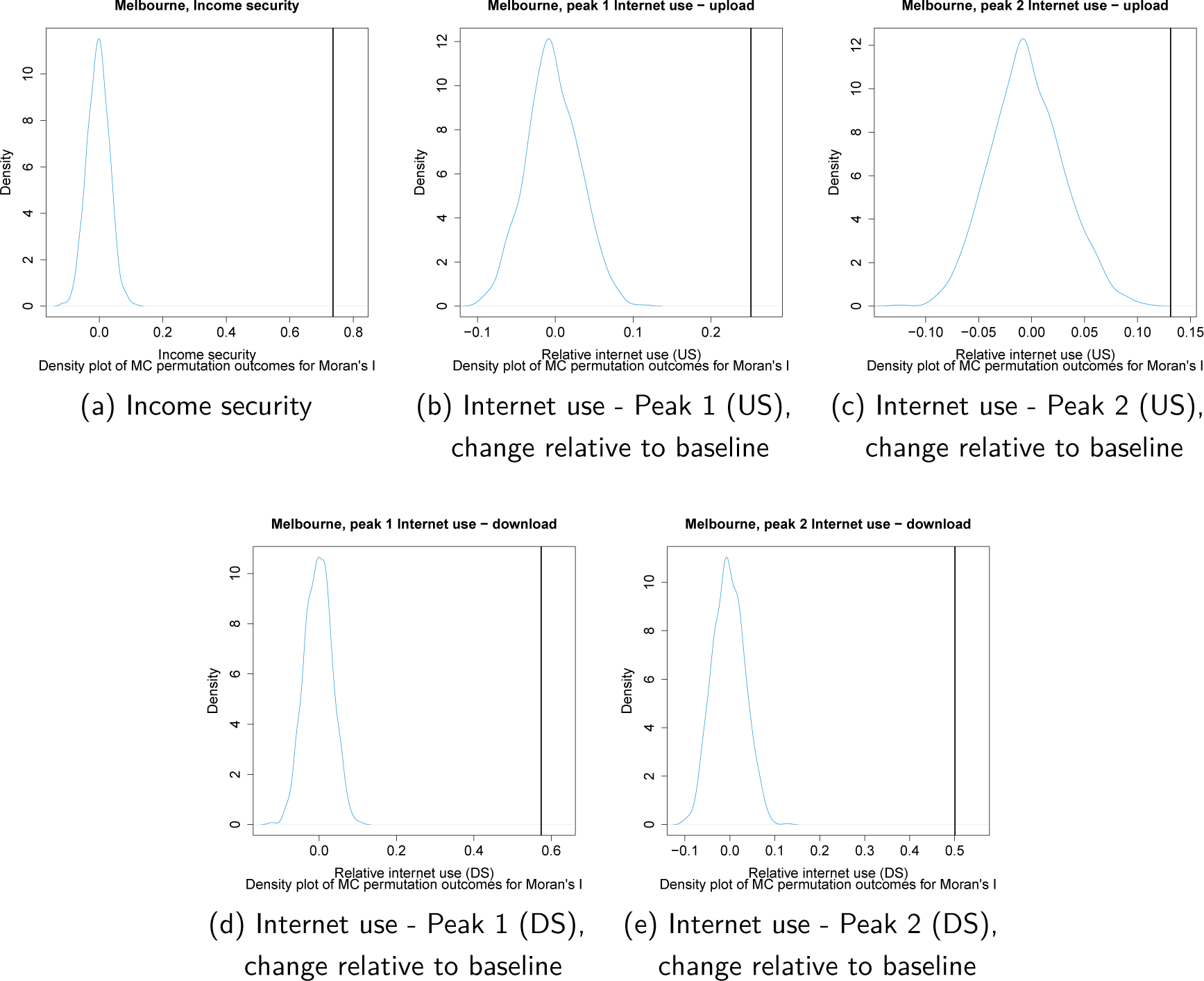
Melbourne - density plots of Moran’s I permutations to complement the tests of statistical significance of Moran’s I statistics for income security (a), as well as relative change to upload volume at peak 1 (b), relative change to upload volume at peak 2 (c), relative change to download volume at peak 1 (d), and relative change to download volume at peak 2 (e).

**FIG. S9.**
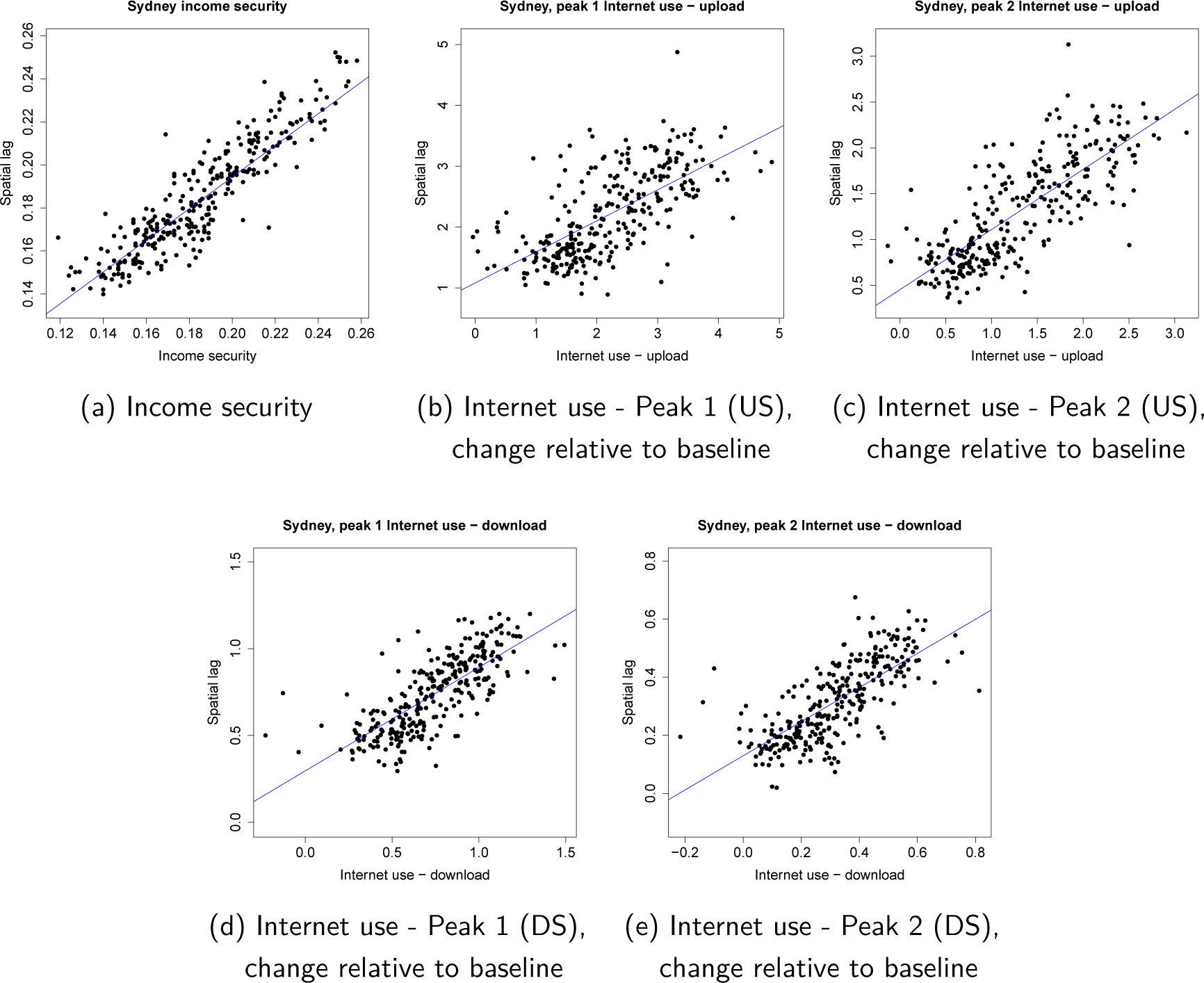
Sydney - Plots of the variables and their spatially lagged values for income security (a), as well as relative change to upload volume at peak 1 (b), relative change to upload volume at peak 2 (c), relative change to download volume at peak 1 (d), and relative change to download volume at peak 2 (e).

**FIG. S10.**
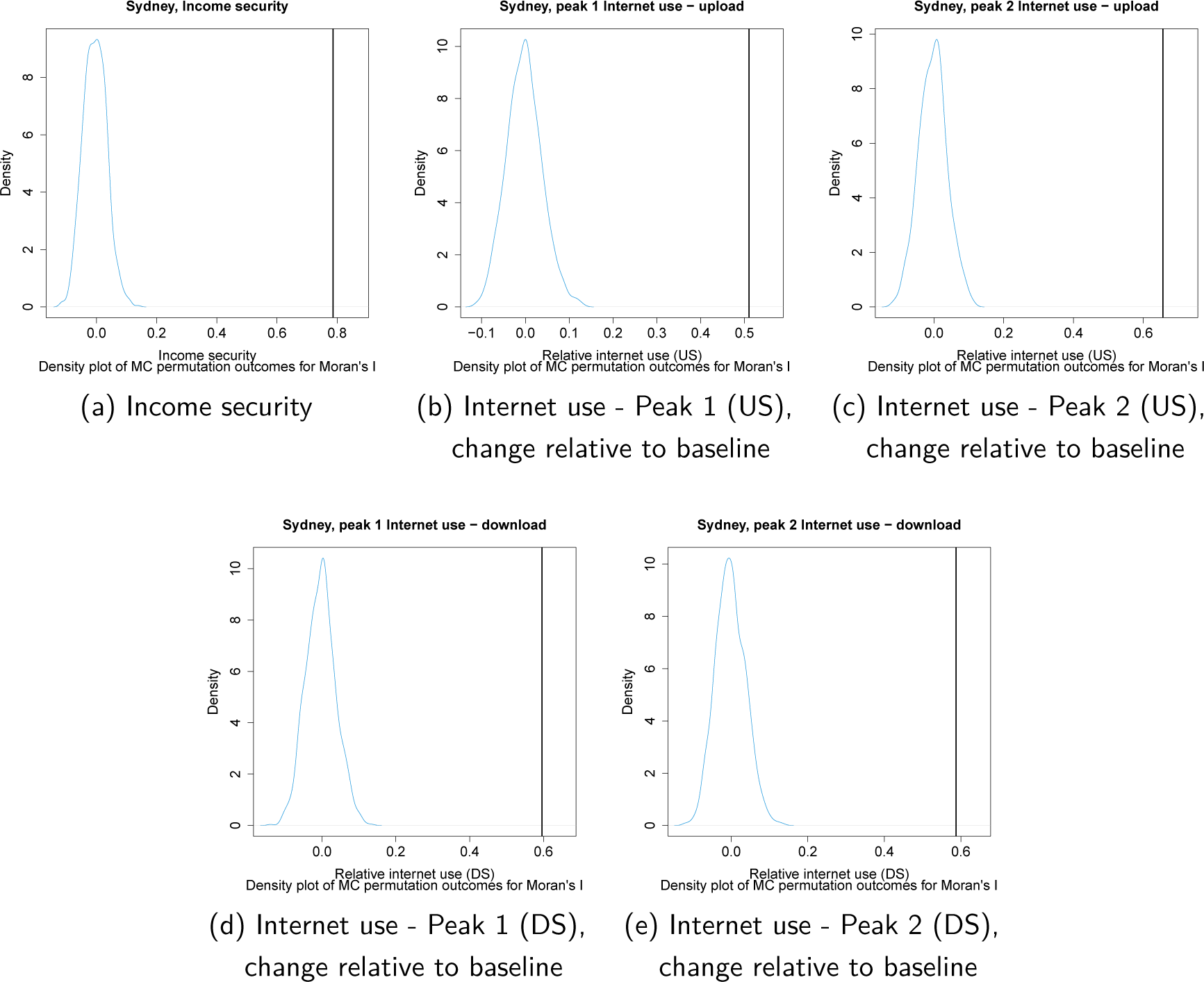
Sydney - Density plots of Moran’s I permutations, complementing tests of statistical significance of Moran’s I statistics for income security (a), as well as relative change to upload volume at peak 1 (b), relative change to upload volume at peak 2 (c), relative change to download volume at peak 1 (d), and relative change to download volume at peak 2 (e).

## Notes

### Competing Interest Statement

The authors have declared no competing interest.

### Funding Statement

Cameron Zachreson, Nicholas Geard, and Martin Tomko were supported by an internal seed grant from the Melbourne School of Engineering, of the University of Melbourne.

### Author Declarations

The CARE study was by approved by the University of Melbourne Human Research Ethics Committee (2056694). The ethics committee approval applied to all study sites.

### Summary of Updates

Altered paper structure to improve clarity. Expanded discussion of study limitations. Added spatial autocorrelation analysis to supplement.

## REFERENCES

1. International Labour Organisation (ILO). COVID-19 and the World of Work: Country Pol- icy Responses. ILO, Geneva, Switzerland; 2020. https://www.ilo.org/global/topics/coronavirus/regional-country/country-responses/lang--en/index.htm.

2. Australian Government, The Treasury. The JobKeeper Payment: Three-month review; 2020.https://treasury.gov.au/publication/jobkeeper-review.

3. Phillips B, Gray M, Biddle N, et al. COVID-19 JobKeeper and JobSeeker impacts on poverty and housing stress under current and alternative economic and policy scenarios. Australian National University, Canberra. 2020.

4. Anderson G, Frank JW, Naylor CD, Wodchis W, Feng P. Using Socioeconomics to Counter Health Disparities Arising from the COVID-19 Pandemic. BMJ. 2020;369.

5. Bambra C, Riordan R, Ford J, Matthews F. The COVID-19 pandemic and health inequalities. J Epidemiol Community Health. 2020;74(11):964–968.

6. Cubrich M. On the frontlines: Protecting low-wage workers during COVID-19, Psychological Trauma: Theory, Research, Practice, and Policy. 2020;12(S1):S186.

7. Fisher J, Languilaire JC, Lawthom R, Nieuwenhuis R, Petts RJ, Runswick-Cole K, et al. Commu- nity, work, and family in times of COVID-19. Community, Work & Family. 2020;23(3):247–252.

8. Sanderson K, Cocker F, et al. Presenteeism: Implications and Health Risks. Australian family physician. 2013;42(4):172.

9. Abedi V, Olulana O, Avula V, Chaudhary D, Khan A, Shahjouei S, et al. Racial, Economic, and Health Inequality and COVID-19 Infection in the United States. Journal of racial and ethnic health disparities. 2020:1–11.

10. Lee K, Sahai H, Baylis P, Greenstone M. Job Loss and Behavioral Change: The Unprecedented Effects of the India Lockdown in Delhi. University of Chicago, Becker Friedman Institute for Economics Working Paper. 2020;(2020-65).

11. Forsythe E, Kahn LB, Lange F, Wiczer D. Labor Demand in the Time of COVID-19: Evidence from Vacancy Postings and UI Claims. Journal of public economics. 2020;189:104238.

12. Cajner T, Crane LD, Decker RA, Grigsby J, Hamins-Puertolas A, Hurst E, et al.. The US labor market during the beginning of the pandemic recession; 2020. National Bureau of Economic Research, https://www.nber.org/papers/w27159.

13. Adams-Prassl A, Boneva T, Golin M, Rauh C. Inequality in the impact of the coronavirus shock: Evidence from real time surveys. Journal of Public Economics. 2020;189:104245.

14. Couch KA, Fairlie RW, Xu H. The Impacts of COVID-19 on Minority Unemployment: First Evidence from April 2020 CPS Microdata. Available at SSRN 3604814. 2020.

15. Montenovo L, Jiang X, Rojas FL, Schmutte IM, Simon KI, Weinberg BA, et al.. Determinants of Disparities in COVID-19 Job Losses; 2020. National Bureau of Economic Research, https://www.nber.org/papers/w27132.

16. Brough R, Freedman M, Phillips D. Understanding socioeconomic disparities in travel behavior during the COVID-19 pandemic. University of California, Irvine Department of Economics Working Paper Series. 2020.

17. Jay J, Bor J, Nsoesie EO, Lipson SK, Jones DK, Galea S, et al. Neighbourhood Income and Physical Distancing During the COVID-19 Pandemic in the United States. Nature Human Behaviour. 2020;4(12):1294–1302.

18. Brynjolfsson E, Horton JJ, Ozimek A, Rock D, Sharma G, TuYe HY. COVID-19 and Remote Work: An Early Look at US Data; 2020. National Bureau of Economic Research, https://www.nber.org/papers/w27344.

19. Bonaccorsi G, Pierri F, Cinelli M, Flori A, Galeazzi A, Porcelli F, et al. Economic and social consequences of human mobility restrictions under COVID-19. Proceedings of the National Academy of Sciences. 2020;117(27):15530–15535.

20. Weill JA, Stigler M, Deschenes O, Springborn MR. Social distancing responses to COVID-19 emergency declarations strongly differentiated by income. Proceedings of the National Academy of Sciences. 2020;117(33):19658–19660.

21. Chiou L, Tucker C. Social Distancing, Internet Access and Inequality; 2020. National Bureau of Economic Research, https://www.nber.org/papers/w26982.

22. Feldmann A, Gasser O, Lichtblau F, Pujol E, Poese I, Dietzel C, et al. The Lockdown Effect: Implications of the COVID-19 Pandemic on Internet Traffic. In: Proceedings of the ACM Internet Measurement Conference; 2020. p. 1–18.

23. Zachreson C, Mitchell L, Lydeamore M, Rebuli N, Tomko M, Geard N. Risk mapping for COVID-19 outbreaks using mobility data. arXiv preprint arXiv:200806193. 2020.

24. Buckee CO, Balsari S, Chan J, Crosas M, Dominici F, Gasser U, et al. Aggregated mobility data could help fight COVID-19. Science (New York, NY). 2020;368(6487):145.

25. Pepe E, Bajardi P, Gauvin L, Privitera F, Lake B, Cattuto C, et al. COVID-19 Outbreak Re- sponse, a Dataset to Assess Mobility Changes in Italy Following National Lockdown. Scientific data. 2020;7(1):1–7.

26. Warren MS, Skillman SW. Mobility changes in response to COVID-19. arXiv preprint arXiv:200314228. 2020.

27. Chang S, Pierson E, Koh PW, Gerardin J, Redbird B, Grusky D, et al. Mobility network models of COVID-19 explain inequities and inform reopening. Nature. 2020:1–6.

28. Australian Statistical Geography Standard (ASGS); 2020. [online; accessed 01- Jun-2021]. https://www.abs.gov.au/websitedbs/D3310114.nsf/home/Australian+Statistical+Geography+Standard+(ASGS).

29. HILDA Survey; 2020. [online; accessed 15-Dec-2020]. https://melbourneinstitute.unimelb.edu.au/hilda.

30. About TableBuilder; 2020. [Online; accessed 12-Aug-2020]. https://www.abs.gov.au/websitedbs/d3310114.nsf/home/about+tablebuilder.

31. Australian Bureau of Statistics Census; 2016. [online; accessed 15-Dec-2020]. https://www.abs.gov.au/census.

32. Sila U, Dugain V. Income, wealth and earnings inequality in Australia: Evidence from the HILDA Survey. OECD; 2019. https://www.oecd-ilibrary.org/economics/income-wealth-and-earnings-inequality-in-australia_cab6789d-en.

33. Shearer FM, Meagher N, Chavez KM, Carpenter L, Pirrone A, Quinn P, et al. Promoting Resilience While Mitigating Disease Transmission: An Australian COVID-19 Study. In: COVID- 19 Pandemic, Geospatial Information, and Community Resilience. CRC Press; 2021. p. 347–362.

34. ABS, 2019, cat. no. 6306.0 - Microdata: Employee Earnings and Hours, Australia; 2019. [online; accessed 01-Sep-2020]. https://www.abs.gov.au/statistics/labour/earnings-and-work-hours/employee-earnings-and-hours-australia/latest-release.

35. LaMontagne AD, Smith PM, Louie AM, Quinlan M, Ostry AS, Shoveller J. Psychosocial and other working conditions: Variation by employment arrangement in a sample of working Australians. American Journal of Industrial Medicine. 2012;55(2):93–106.

36. Louie A, Ostry A, Quinlan M, Keegel T, Shoveller J, LaMontagne A. Empirical study of employment arrangements and precariousness in Australia. Relations Industrielles/Industrial Relations. 2006;61(3):465–489.

37. Dingel JI, Neiman B. How many jobs can be done at home? Journal of Public Economics. 2020;189:104235.

38. Atchison CJ, Bowman L, Vrinten C, Redd R, Pristera P, Eaton JW, et al. Perceptions and behavioural responses of the general public during the COVID-19 pandemic: A cross-sectional survey of UK Adults. medRxiv. 2020.

39. Karaye IM, Horney JA. The impact of social vulnerability on COVID-19 in the US: an analysis of spatially varying relationships. American Journal of Preventive Medicine. 2020;59(3):317–325.

40. Hankivsky O, Kapilashrami A. Beyond sex and gender analysis: an intersectional view of the COVID-19 pandemic outbreak and response; 2020. Gender and Women’s Health Unit, Cen- tre for Health Equity, Melbourne School of Population and Health Equity, https://mspgh.unimelb.edu.au/data/assets/pdf_file/0011/3334889/Policy-brief_v3.pdf.

41. Chung H, Seo H, Forbes S, Birkett H. Working from home during the COVID-19 lockdown: Changing preferences and the future of work; 2020. Kent Academic Repository, University of Kent, https://kar.kent.ac.uk/83896/1/Working_from_home_COVID-19_lockdown. pdf.

42. de Haas M, Faber R, Hamersma M. How COVID-19 and the Dutch ‘intelligent lockdown’ change activities, work and travel behaviour: Evidence from longitudinal data in the Netherlands. Transportation Research Interdisciplinary Perspectives. 2020;6:100150.

43. Bivand RS, Pebesma E, Gomez-Rubio V. Applied spatial data analysis with R, Second edition. Springer, NY; 2013. Available from: https://asdar-book.org/.

44. Bivand R, Wong DWS. Comparing implementations of global and local indicators of spa- tial association. TEST. 2018;27(3):716–748. Available from: https://doi.org/10.1007/s11749-018-0599-x.

